# Predicting undiscovered non-human primate hosts of Semliki Forest complex Alphaviruses

**DOI:** 10.64898/2026.08.10.26360069

**Authors:** Michael Celone, Adrian Castellanos, Bernard Okech, Sean Beeman, Simon Pollett, Barbara Han

## Abstract

Arthropod-borne *Alphaviruses* in the Semliki Forest (SF) virus complex, including Chikungunya virus, Mayaro virus, and O’nyong-nyong virus, represent a substantial threat to human health globally. These antigenically related viruses often cause short-term febrile symptoms that can progress to chronic and debilitating arthropathy. The ecology of these viruses is complex due to the involvement of various animal hosts and mosquito vectors in their transmission cycles. Non-human primates (NHPs) have been identified as potentially important animal hosts that may contribute to ongoing transmission and emergence, but the full range of known NHP hosts is not clear. Due to the epidemiological importance of NHPs, we predicted NHP species with a high probability of being carriers of SF complex *Alphaviruses*. We first compiled an extensive database of intrinsic and extrinsic NHP traits including reproduction, diet, behavior, biogeography, home-range, and climate. Next, we identified NHP species that are known zoonotic hosts of SF complex *Alphaviruses.* Hosts are defined as naturally infected NHPs identified through field studies. They do not necessarily meet the criteria for reservoir competence. Host vs. non-host status was largely determined through serology and species without data were treated as non-hosts in our analysis. Finally, we used boosted regression trees (BRT) to develop a trait profile of the known NHP host species. Using this trait profile, we identified additional, potentially unrecognized NHP hosts with a comparable trait profile. We found that latitudinal range, maximum longevity, maximum temperature, minimum human population density, number of ecoregions in species range, neonate mass, female mass, and mean precipitation were important predictors of zoonotic host status. Additionally, we were able to distinguish NHP hosts from non-hosts, and to identify 30 additional NHP species predicted to carry SF complex *Alphaviruses*. These findings can serve as hypotheses that can guide targeted surveillance and may help direct additional field epidemiological studies to better define the risk and risk factors of *Alphavirus* emergence.

**Author Summary:** *Alphaviruses* including Chikungunya virus, Mayaro virus, and O’nyong-nyong virus, represent a major global health threat. Non-human primates are potentially important animal hosts in the transmission and emergence of *Alphaviruses*, but the ecology of these viruses is still poorly understood. Due to the importance of NHPs in *Alphavirus* transmission, we predicted non-human primate species with a high probability of being carriers of *Alphaviruses*. We compiled a database of non-human primate traits such as reproduction, diet, and behavior and then identified the trait profile of non-human primate species that are known zoonotic hosts of *Alphaviruses.* We found that traits such as latitudinal range, maximum longevity, and maximum temperature were important predictors of zoonotic host status. Finally, we identified additional, potentially unrecognized non-human primate hosts with a comparable trait profile. These findings can guide targeted surveillance and may help direct additional field studies to better define the risk of *Alphavirus* emergence. By prioritizing certain non-human primate species for future surveillance, our findings may improve the efficiency of wildlife sampling and enhance early detection of zoonotic *Alphaviruses*.

## Introduction

*Alphavirus* is a genus of enveloped positive-stranded RNA viruses from the family Togaviridae. Several *Alphaviruses*, including Chikungunya virus (CHIKV) and Mayaro virus (MAYV) have caused major outbreaks in human populations [1, 2]. One phenotypic classification of *Alphaviruses* is whether they present with either encephalitic or arthritogenic clinical manifestations [3]. The arthritogenic *Alphaviruses* may cause nonspecific febrile illness with rash, headache, fever, and arthralgia, but chronic joint or muscle pain can persist for many months after infection [4, 5]. The neotropical urban-cycle CHIKV epidemic and ongoing global burden of endemic infections has prompted concern about the pandemic potential of other arthritogenic *Alphaviruses*, for example MAYV [4, 6, 7].

The Semliki Forest (SF) virus complex is a group of antigenically related *Alphaviruses* that cause similar arthritogenic sequelae [8]. Human viruses in the SF complex include MAYV, CHIKV, Semliki Forest virus (SFV), O’nyong-nyong virus (ONNV), Una virus (UNAV), and Ross River virus (RRV) [8]. Although CHIKV has a global distribution, the other viruses are limited to specific geographic regions. For example, MAYV occurs in Latin America [9], RRV occurs in Australia and other islands of the South Pacific [10], and sporadic ONNV outbreaks have occurred in several countries throughout sub-Saharan Africa [11]. Although the ecology of the SF complex viruses is still poorly understood, all are transmitted between mosquito vectors and non-human vertebrate animals in a sylvatic cycle, with occasional outbreaks in human populations [4, 5] and, in the case of CHIKV, an ongoing widespread urban transmission cycle. Non-human primates (NHPs) have been identified as a potential host for several viruses in the SF complex based on viral isolates obtained from infected NHPs as well as serological evidence of past infection [12, 13].

Many emerging zoonotic viruses of global significance have spilled over from animal hosts into human populations [14]. Identifying the non-human animal populations that serve as disease hosts can provide valuable insight into the ecology, transmission, and geographic distribution of viruses and may support risk prediction efforts. Well-defined animal hosts can serve as sentinel populations that are monitored for early outbreak detection and can aid in forecasting pathogen spillover events [15]; for example, captive and free-ranging birds for West Nile Virus surveillance [16] and non-human primates for yellow fever virus surveillance [17]. Ongoing disease surveillance is especially important given that anthropogenic activities and habitat fragmentation may increase interaction between humans and wild animals such as NHP species that occupy urban forests in Brazil [15]. Identification of competent animal hosts is a vital step in reducing the risk of transmission at the human-animal interface. In addition, identification of non-human hosts is important because the maintenance of a pathogen in a non-human animal population can complicate local disease control efforts (e.g., sylvatic yellow fever outbreaks in Brazil [18]).

Because of the difficulties associated with disease detection in wild-caught animal populations (including the large number of potential species which may be undiscovered hosts), trait-based machine learning has emerged as a promising way to predict animal species that may serve as disease hosts, enabling more focused, confirmatory field studies. This method uses certain life history, ecological, biogeographical, or physiological traits (e.g., body mass, sexual maturity age, or growth rate) *a.k.a* “trait profile” to distinguish disease hosts from non-hosts. The “trait profile” of known pathogen hosts can be used to calculate a probability of host status for other species based on similarity of trait profiles. This method has been used previously to identify high-probability filovirus carriers which were later confirmed in field studies. For example, a field study published in 2017 [19] confirmed *Eonycteris spelaea* bats as a novel host of filoviruses after they were previously identified as potential filovirus-positive on the basis of “trait profile” similarity with known filovirus-positive bats. Since our knowledge of the *Alphavirus* host range remains incomplete, this study attempted to assess additional NHP host status for human *Alphaviruses* in the SF virus complex. We sought to develop a trait profile of the known NHP carriers of SF complex viruses and to identify other NHP species with a similar trait profile. The results of this analysis will clarify the complex ecology of these important viruses and guide future ecological studies and disease surveillance efforts.

## Methods

### Outcome Data

We first reviewed the Global Virome in One Network (VIRION), an atlas of vertebrate-virus associations compiled by the Verena Consortium [20], to identify known NHP hosts of wild-caught SF complex *Alphaviruses*. In the current study, host status was based on detection of the pathogen in a wild-caught animal, rather than full criteria for reservoir status [21]. Results from experimental animal model studies were excluded because they do not reflect natural transmission dynamics and we intended to identify NHP species that serve as true hosts in natural settings. We cross-checked this dataset against NHP *Alphavirus* positivity that was documented in the GIDEON database [22]. This dataset was further supplemented with several additional NHP species that were identified as MAYV or CHIKV carriers in two recent systematic reviews [12, 13]. Finally, we conducted a search of PubMed in December 2022 to determine if we had missed any additional studies that reported NHP *Alphavirus* host status. We used the following search terms (without language restrictions or date limits): *(primate* OR monkey*) AND (alphavirus OR “Chikungunya virus” OR “Mayaro virus” OR “Una virus” OR “Ross River virus” OR “O’nyong-nyong virus” OR “Semliki forest virus”)*.

We followed the methods used in previously published trait-based ecological modeling studies [23–25] to identify carriers and non-carriers of SF *Alphaviruses*. Each NHP species in our dataset was assigned a binary label: 1 = carrier (host) of at least one virus from the SF *Alphavirus* complex; or 0 = not known to be a carrier (non-host). For this study, “carriers (hosts)” were defined as any NHP species with evidence of natural, wild-caught infection by SF complex *Alphaviruses* via diagnostic techniques including serology, PCR, or isolation of live virus. We excluded evidence of infection from experimental laboratory studies, focusing only on wild-caught infections. We treated species that were reported to be seropositive as carrier (host) = 1, even if there were no additional diagnostics to confirm active or recent infection for that species. When experimental inoculation data is not available, repeated surveys and large sample sizes are necessary to establish that a given species is not able to harbor an infection. Thus, we used a conservative definition of host status for this analysis, whereby “unknown” hosts were considered non-hosts. Given pervasive lack of surveillance for *Alphaviruses* and similarly under-surveilled zoonotic pathogens in wildlife, we selected these label designations to achieve a balance between false positive versus false negative predictions in our study. This binary split enables the model to learn the traits of confirmed and suspected host species to support the prediction of potential carriers from a larger pool of potential host species for which no surveillance data exist. Ideally, the baseline classification performance of our model will improve as more NHP hosts are identified in future studies. A list of NHP carriers, virus detection methods, and citing literature is provided in S1 Table.

### Trait Data

Next, we compiled NHP trait data from several sources, including the COMBINE mammal database of intrinsic and extrinsic traits [26] and the Ecological Traits of the World’s Primates [27]. The Coalesced Mammal Database of Intrinsic and Extrinsic Traits (COMBINE) dataset contains 54 trait variables for more than 6,000 extant and recently extinct mammal species. This data includes intrinsic and extrinsic traits related to morphology, reproduction, diet, biogeography, life-habit, phenology, behavior, home range and density. Twenty-seven non-biogeographical traits were imputed phylogenetically by the COMBINE authors using the *missForest* algorithm if greater than 20% of data was available [26]. The Ecological Traits of the World’s Primates database contains data on home range, population trend, conservation status, locomotion, trophic guild, diel activity, mass, habitat, and geographic realm for 504 NHP species. Data is derived from 1,216 studies published between 1941 and 2018 [27].

We also developed several variables from additional datasets in order to capture the impact of climate, biogeography, and human influence on NHP host status. We first downloaded area of habitat maps for all NHP species, which represent the suitable areas for each species within its geographic range [28]. These area of habitat maps were then used to calculate the following variables: mean, minimum, and maximum temperature and precipitation using the WorldClim bioclimatic variables [29]; mean, minimum, and maximum human population density using the Gridded Population of the World (GPW), version 4 [30]; mean mammal species richness using the 2024 IUCN mammal species richness raster [31]; mean “human footprint” (i.e., a composite measure of human pressure on the environment that incorporates different variables such as population density, crop lands, roads, etc.) using the Human Footprint 2018 release [32]; terrestrial ecoregions of the world [33]; and several measures of species geographic range size [34].

In order to capture taxonomic clustering of SF complex host status we also included primate family as a series of binary variables in the analysis. Finally, we calculated several additional variables that were derived from variables in the COMBINE dataset, following previous studies that suggest that traits indicating life history pace distinguish zoonotic hosts from non-host species [23]. These derived variables included postnatal growth rate (weaning body mass divided by neonatal body mass), mass specific production (based on the formula from [35]), home range scaled by adult mass, relative age at sexual maturity (defined as age at sexual maturity divided by maximum longevity), relative age at first birth (defined as the age at first birth divided by maximum longevity), and body size ratio (defined as adult body mass divided by neonate body mass).

After merging the datasets described above, we removed variables with more than 80% missing values or those with zero variance among the NHP species. The full list of traits is presented in S2 Table. The final dataset consisted of 116 features across 505 NHP species. Twenty-five primate species were removed from the database due to complete missingness. In addition, the COMBINE dataset included eight NHP species that were recently reclassified as sub-species. These eight sub-species were not included in the area of habitat maps mentioned above, and therefore, were dropped from the present analysis.

## Data Analysis

Machine learning algorithms, especially boosted regression trees (BRTs), have been employed with high accuracy to predict unidentified animal hosts of zoonotic pathogens such as Ebola virus and Nipah virus [23, 24, 36, 37] as well as undiscovered arthropod vectors of disease [38–41] based on species-level traits. A recent modeling study that compared several methods to predict likely hosts of severe acute respiratory syndrome coronavirus-2 (SARS-CoV-2) found that BRTs outperformed other modeling strategies including network models and Bayesian additive regression trees [42, 43]. Machine learning algorithms like BRTs are well-suited for ecological data because they can accommodate different types of predictors, model complex and nonlinear relationships between predictors and an outcome, and do not require assumptions about the underlying distribution of the data [44]. These methods may also be less susceptible to some sampling biases mentioned above such as unequal survey effort or research capacity between countries [23].

Using the *gbm* package (Version 2.1.8.1) in the R software (Version 4.2.2) [45], we trained a BRT model on the 505 extant NHP species in our dataset. Code and data for this analysis are available at: https://github.com/mike-celone/NHP_alphavirus. The model was trained on 80% of the dataset with the remaining 20% held out for testing. We implemented the 80/20 train-test split using stratified random sampling on the binary outcome variable and repeated this procedure across bootstrap runs with performance metrics averaged over iterations. We specified a Bernoulli-distributed error for our binary response variable (host vs. non-host) and used 5-fold cross-validation during model training to prevent over-fitting and permutation procedures to generate variable importance scores. Optimal hyperparameters for the model were determined using a grid search.

To further assess the statistical accuracy of our model, we used a “target shuffling” approach that has been described in previous trait-based modeling studies [24, 36, 41]. Target shuffling is a technique to evaluate if a model is overfitting to data and detecting misleading or incorrect correlations by chance alone [46]. We first performed a bootstrapped permutation analysis that involved randomly shuffling the binary outcome labels to calculate a null AUC across 10 model iterations. The AUC for the null model should ideally be AUC = 0.50; otherwise, the model is overfitting to the data. We used this null AUC to calculate a corrected AUC for our final BRT model, defined as:

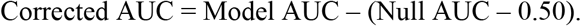

### Study Effort

We also explored the potential impact of unequal study effort on our model results. The goal of this analysis was to determine if our model was truly distinguishing SF complex *Alphavirus* hosts from non-hosts or was merely identifying the most studied NHP species. Following previous trait-based analyses [23, 24, 36, 37], we used Web of Science (WoS) citation count as a proxy for study effort. We manually searched the WoS using each NHP species name as a search string and compiled the total citation count for each NHP species. We then conducted a second BRT analysis using WoS citation count as the outcome variable with a Poisson-distributed error term, and the same set of feature variables for each species. We followed the same grid search procedure described above to identify the optimal hyperparameters for this model. Finally, we compared the trait profile of the most studied species with the trait profile of SF complex *Alphavirus* hosts to assess the impact of “studiedness” on model results. The results of this citation prediction model are presented in the Supplementary Materials.

#### Variable Selection

We omitted variables with relative importance scores <1% in order to ensure that our model was parsimonious and more easily interpretable and to avoid overfitting. We calculated the relative percent contribution for each variable as a measure of variable importance. This metric quantifies how often the model selects a variable for splitting and weights the scores by the squared improvement to model performance averaged across all trees [44]. We first ran the model with all variables included (except for variables that were omitted because of >80% missing values or those with zero variance). We subsequently re-fit the model using only variables with a relative percent contribution greater than 1%. The main model presented in this paper includes the 30 variables with >1% relative contribution (see Results). Eighty-four variables from the initial dataset were dropped (see Results).

### Mapping hotsposts

We mapped the geographic range of known SF complex hosts and potential hosts. We identified potential hosts using the predicted probability value that maximized true positive and true negative NHP species. Maps represent the overlapping geographic ranges of predicted host species using area of habitat rasters mentioned previously. Grid-cell-level hotspot maps were generated by summing the number of predicted host species whose area of habitat overlapped each cell. All spatial analyses were conducted in a common geographic coordinate reference system (WGS84; EPSG:4326). These maps were created using the area of habitat maps mentioned previously.

## Results

### Known NHP hosts and associated traits

We identified 52 NHP species from the literature that were identified as SF complex *Alphavirus* hosts. These included 35 hosts of CHIKV, 16 hosts of MAYV, 14 hosts of ONNV, nine hosts of SFV, and one host of UNAV. The majority of known NHP hosts were from the family Cercopithecidae (n=29 species) and resided in sub-Saharan Africa and Southeast Asia (S1 Table). Serological tests were most commonly used to identify known hosts although live virus was isolated from eight known host species (S1 Table). Of the 52 species, 44 were identified by serology alone, three by virologic methods alone, and five by both.

Our model demonstrated a relatively strong performance with a corrected AUC of 0.91. According to the relative importance scores, the variables that were most important in distinguishing known hosts from non-hosts included latitudinal range, maximum longevity, maximum temperature, minimum human population density, number of ecoregions with the area of habitat, neonate mass and female mass. The full list of variables and importance scores are included in S4 Table. Partial dependence plots (Fig 1) revealed that SF complex *Alphaviruses* tend to be found in NHP species with a large latitudinal range in hotter regions, that are relatively long-lived and social with large home ranges containing diverse ecoregions.

**Fig 1.**
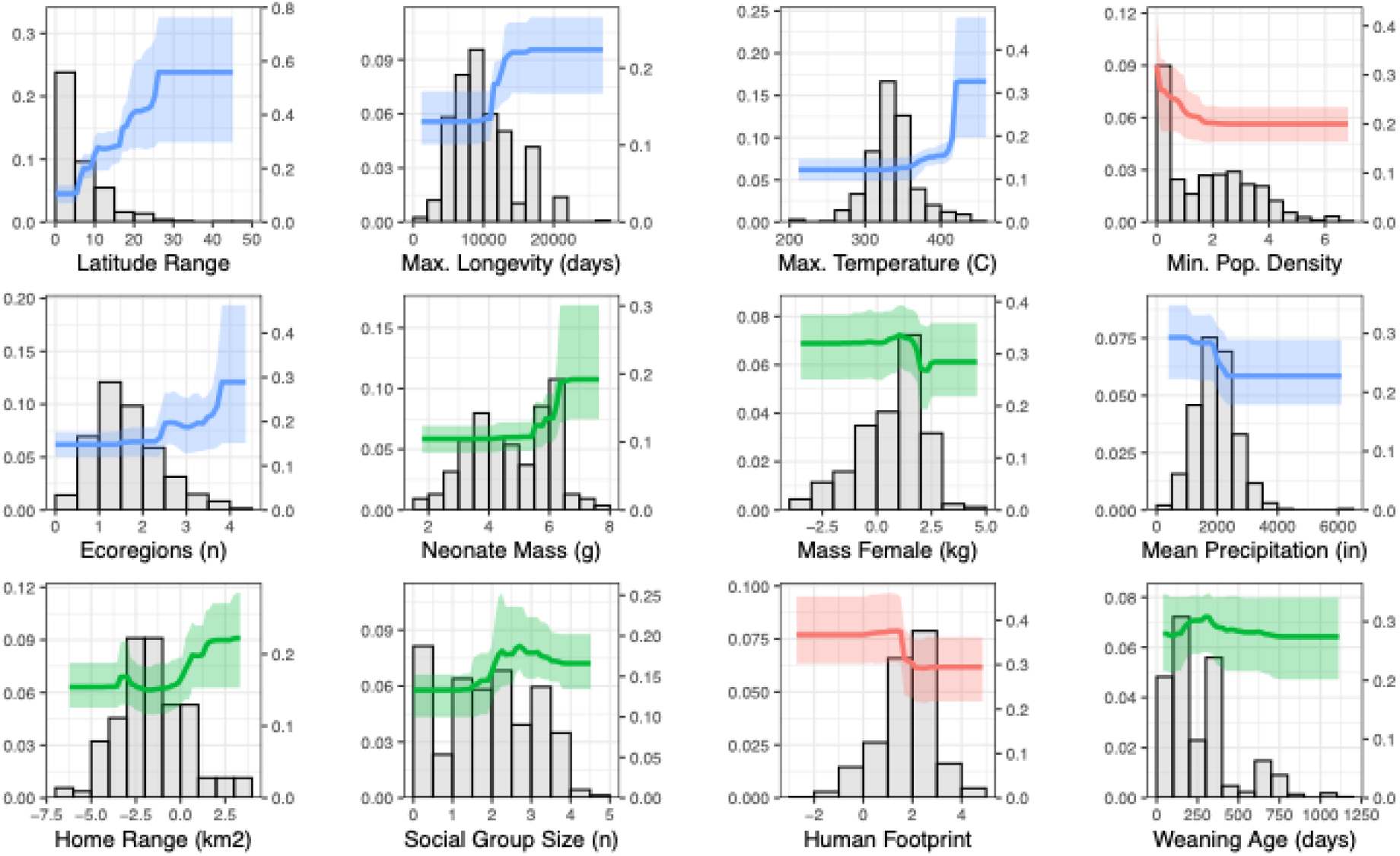
The trait profile of SF Alphavirus positive NHP species. Partial dependence plots of the top 12 predictor variables from the BRT analysis illustrate the trait profile of NHP species that are known zoonotic hosts of SF complex *Alphaviruses*. Traits are displayed in descending order of relative importance for distinguishing hosts from non-hosts, arranged from left to right and top to bottom, and color coded according to type (human influence in red, life history in green, and biogeography in blue). Frequency histograms illustrate the distribution of trait values across all NHP species. Bold lines represent the mean value calculated from 10 bootstrap iterations, while shaded areas indicate the 95% confidence interval. On the left y-axis, we depict the marginal impact of each variable (x-axis) on model prediction accuracy. The right y-axis represents the proportion of species falling within each histogram bar (bin). Ecoregions, mass female, home range, and human footprint variables have been log-transformed. Units for additional variables include the following: temperature = °C x 100; longevity = days; mass = g; latitude = decimal degrees; population density = persons/km2.

### Predicted NHP hosts

Based on the trait profile of known SF complex *Alphavirus* hosts, we sought to identify undiscovered NHP hosts of SF complex *Alphaviruses*. The predicted probability value that maximized true positive and true negative NHP species was 0.122. Using this value, we identified 30 additional NHP species predicted to carry SF complex *Alphaviruses*. The five undiscovered NHP species with the highest predicted probability (*Chlorocebus tantalus, Papio cynocephalus*, *Chlorocebus cynosures, Colobus angolensis,* and *Papio kindae*) have a wide distribution across East, Central and West Africa. We also identified several NHP species in South America with high predicted probability, including the common wooly monkey (*Lagothrix lagothricha*), Peruvian spider monkey (*Ateles chamek*), Black capuchin (*Sapajus nigritus*), and the red-faced spider monkey (*Ateles paniscus*). These species are found in Brazil, Peru, Suriname, Guyana, and French Guiana. The complete list of NHP species and the predicted probability associated with each species is presented in S5 Table.

The trait profile of the most studied species (S3 Table) was markedly different from the trait profile of predicted hosts, indicating that the model was successful at distinguishing SF complex *Alphavirus* hosts from non-hosts and not merely identifying the most studied NHP species. The most important variables in this analysis included number of ecoregions, maximum population density, dispersal, altitude breadth, and maximum latitude.

### Geographic range of known and predicted hosts

Overlapping geographic range maps for the known and predicted NHP hosts of SF complex *Alphaviruses* are presented in Figs 2 and 3. The highest concentration of overlapping known and predicted host species is found in Central-West Africa, especially in Gabon and the Democratic Republic of Congo. The inclusion of predicted host species in Fig 3 reveals additional locations with multiple overlapping NHP species such as Angola, eastern Peru, southern portions of Venezuela and Colombia, and western Brazil.

**Fig 2.**
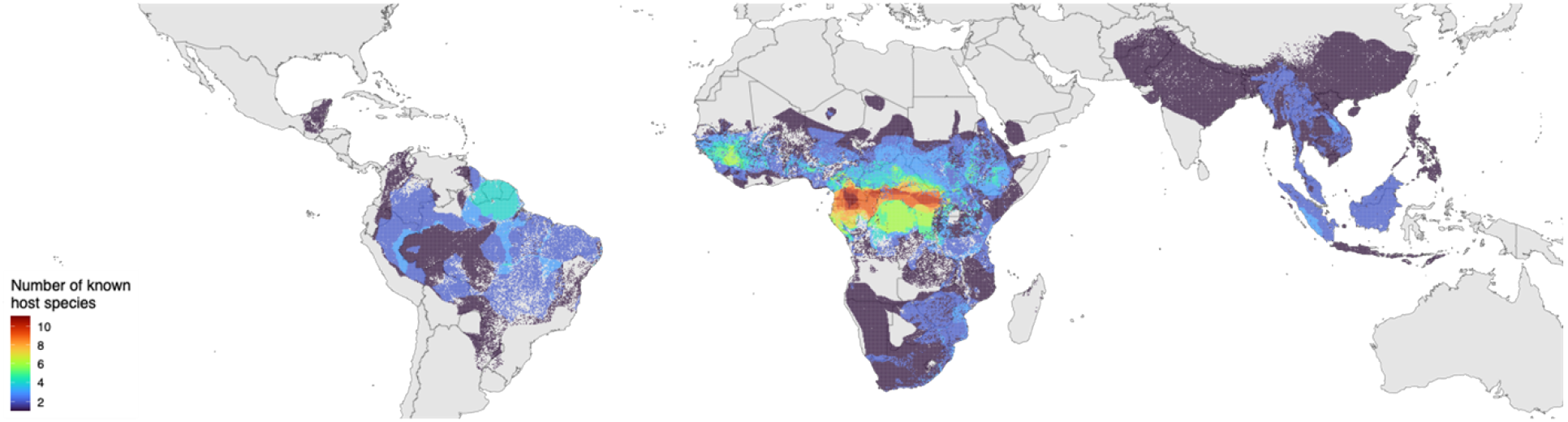
Geographic range maps of known SF complex *Alphavirus* host NHP species. Overlapping geographic ranges of non-human primate (NHP) species with documented evidence of natural infection with SF complex *Alphaviruses*. Maps were created in R using shape files from the Natural Earth public domain repository (http://www.naturalearthdata.com/).

**Fig 3.**
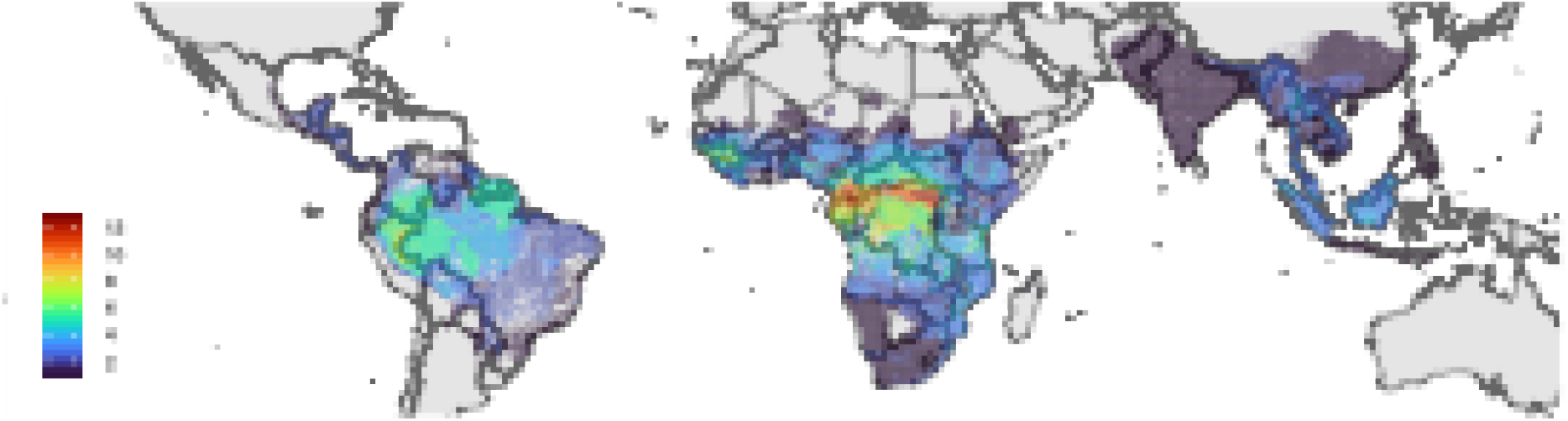
Geographic range maps of known and predicted SF complex *Alphavirus* host NHP species. Overlapping geographic ranges of NHP species with documented evidence of natural infection with SF complex *Alphaviruses* and species predicted by the boosted regression tree (BRT) model to be potential hosts. Maps were created in R using shape files from the Natural Earth public domain repository (http://www.naturalearthdata.com/).

## Discussion

In this study we present an analysis of the known and predicted NHP carriers of SF complex *Alphaviruses*. This study provides an important addition to the literature regarding the ecology of *Alphaviruses*, an increasingly important genus of viruses with outbreak, epidemic, and pandemic potential. Due to a relative paucity of studies regarding the ecology and potential non-human reservoirs of *Alphaviruses*, we currently do not have a complete understanding of the *Alphavirus* host range. An important step in the surveillance of these viruses is a better understanding of predictors of positivity in wildlife (whether this is seropositivity or isolated live virus). Due to the paucity of data in other animal species, and the potential importance of NHPs in several *Alpahvirus* transmission systems, we decided to focus our current analysis on NHPs.

The identification of animal hosts is difficult for a number of logistical reasons. For example, researchers may face challenges such as accessing animal populations in remote locations, obtaining a sufficient number of samples for detecting disease, the difficulty and costs associated with complex wildlife trapping procedures, and the availability and cost of obtaining sampling materials and storing/shipping samples [47]. There are also many limitations related to diagnostics that can impede the identification of animal hosts. For instance, cross-reacting antigens of closely related viruses can compromise diagnostic test results and tests designed for domestic animals may have limited accuracy in wild populations [47].

Machine learning algorithms have been employed with high accuracy to predict unidentified animal hosts of zoonotic pathogens [23, 24, 36, 37] as well as undiscovered arthropod vectors of disease [38, 39] based on species-level traits. These methods are particularly useful because they are less susceptible to the sampling biases mentioned above (e.g., unequal survey effort or research capacity between countries) [23]. Ecological trait-based models were validated using real-time data during the SARS-CoV-2 pandemic and outperformed other models in predicting potential bat hosts of betacoronaviruses [42]. Furthermore, trait-based predictions of undiscovered filovirus hosts [24] were validated in several independent studies of wild-caught bats [19, 48].

Using a comprehensive set of covariates spanning several domains (e.g., life history, biogeography, human influence, etc.) we developed a trait profile of known SF complex *Alphavirus* NHP hosts. The covariates with the greatest relative importance included latitude range, temperature, human population density, number of ecoregions, longevity, and neonate mass. Based on this trait profile, we identified additional NHP species with a high probability of being SF complex *Alphavirus* hosts, including NHP species in both Africa (*Papio cynocephalus*, *Papio kindae*, *Chlorocebus cynosures*, and *Chlorocebus tantalus)* and South America (*Lagothrix cana* and *Ateles paniscus*). The model distinguished hosts from non-hosts with relatively high accuracy and the results did not appear to be particularly biased by the impact of unequal study effort. The results of this analysis can provide useful information to epidemiologists or ecologists who are interested in the geographic distribution or the transmission cycle of *Alphaviruses* and can prompt targeted fieldwork. Clarifying the *Alphavirus* host range is an important step in understanding the ecology of these important viruses and determining where human populations may be at risk of disease spillover.

The NHP species identified in this study as high-probability carriers can serve as a starting point for future studies (either field-based surveillance or laboratory) focused on the ecology of SF-complex *Alphaviruses*. The geographic range of several of these NHP species overlap with high-density human settlements, increasing the potential for virus transmission between humans and primates. According to the IUCN RedList, 33% of the predicted additional hosts identified by our model are found in some form of human modified habitat, such as plantations, urban areas, rural gardens, and heavily degraded forests [49]. These species are good candidates for further studies aimed at clarifying the disease ecology and the precise role of various NHP species in virus transmission. For example, the geographic range of the *Chlorocebus tantalus* species overlaps several densely populated urban areas in West and Central Africa including in Ghana and Nigeria. Furthermore, this species is highly adaptable to different environments and is able to live in various habitats including in cities. *Papio cynocephalus* is another NHP species that is widely distributed across countries with high human population in East and Central Africa. This species also exhibits behaviors that bring them into close proximity with human settlements, such as garbage-foraging [50]. Therefore, they could be targeted for additional study to clarify their competence as virus hosts.

The traits that we identified as important for distinguishing carriers vs. non-carriers were similar to another study focused on the potential hosts of *Flaviviruses [25]*. For example, traits related to geographic range size, maximum longevity, and neonate body mass emerged as important variables in both studies. There were also several known *Alphavirus* carriers identified as high probability *Flavivirus* carriers in the previous study, including *Papio anubis* and *Cercopithecus mitis*, as well as several NHP species that are known to be carriers of both *Alphavirus* and *Flavivirus*, including *Macaca mulatta*, *Chlorocebus aethiops*, and *Macaca fascicularis*, among others. We also identified human population density as an additional important predictor of *Alphavirus* host status, highlighting the importance of geographic overlap between *Alphavirus* carriers and populated human settlements. Human encroachment into the natural habitats of these species through deforestation or hunting leads to increased interaction between NHPs and human populations [51]. Furthermore, fragmentation of NHP habitats leads to behaviors such as garbage foraging and crop-raiding, that may lead to the presence of NHPs in urban environments [52]. This dynamic is an important consideration for public health professionals concerned with disease transmission between NHPs and human populations.

Although this study provides an important foundation for further research into *Alphavirus* ecology, there are several important limitations that should be considered. While we attempted to assess the impact of studiedness on our model, it is likely that under-sampling of less studied species limits our understanding of the true *Alphavirus* host distribution. Ideally, the results of our study and similar modeling studies can guide field surveillance efforts and assist ecologists in identifying those under-sampled NHP species that can be targeted for further study.

An additional limitation is the use of serology to assess host status, especially when samples are collected at a single time point [53]. Serological assays can exhibit cross-reactivity among closely related viruses, potentially leading to misclassification of host status [54]. However, serology represents a major source of evidence for wildlife exposure to pathogens, and inclusion of these data allows for broader characterization of the potential NHP host species of *Alphaviruses*. Furthermore, our analysis includes a group of closely related *Alphaviruses*, rather than a single virus. Because we did not focus on virus-specific interpretation, any cross-reactivity and potential misclassification of host status is unlikely to have a major impact on our conclusions.

Data sparsity is another significant limitation of ecological modeling studies [25]. Although BRT models are adept at handling missing data, our understanding of life history, behavior, biogeographical, and other traits of under-studied NHP species is limited. Therefore, the results of our study should be interpreted cautiously, in the context of the available information. As additional NHP trait data is collected in the future, our ability to make predictions about likely disease reservoirs will improve. One promising avenue of future research could involve mining life history data for certain traits (e.g., litter size, mass, etc.) from natural history collections. This method of data collection would be easier and less costly than field-based studies, especially for NHP species that are not easily accessible. For example, digitized museum specimens were used to study the impact of climate and body size on litter size in small mammals [55].

In addition, the results of our study are meant to serve as an initial evidence base for future research into *Alphavirus* ecology. However, identifying a true animal “reservoir” is a complex process that involves both laboratory and field based surveillance studies [56]. Therefore, the identification of high-probability hosts in the current study does not imply that these species are necessarily competent *Alphavirus* reservoirs, but should instead provide guidance for future studies aimed at improving the existing evidence base related to *Alphavirus* ecology. Furthermore, we did not include mosquito vector distribution data in the model. Potential host species were identified solely based on ecological and life-history traits, so the model cannot directly infer actual transmission cycles. Predicted host hotspots represent potential exposure to *Alphaviruses* rather than actual transmission risk, which additionally depends on the presence and abundance of mosquito vectors.

### DISCLAIMER

The contents, views or opinions expressed in this publication are those of the authors and do not necessarily reflect official policy or position of Henry M. Jackson Foundation for the Advancement of Military Medicine, Inc., Uniformed Services University of the Health Sciences, the Department of War (DoW), or Departments of the Army, Navy, or Air Force. Mention of trade names, commercial products, or organizations does not imply endorsement by the U.S. Government.

## Data Availability

The minimal data set and accompanying code is available at Github via https://github.com/mike-celone/NHP_alphavirus

## ACKNOWLEDGEMENTS

This project was supported by the Verena data ecosystem, funded by the U.S. National Science Foundation (NSF DBI 2213854).

---

**Table S1.**
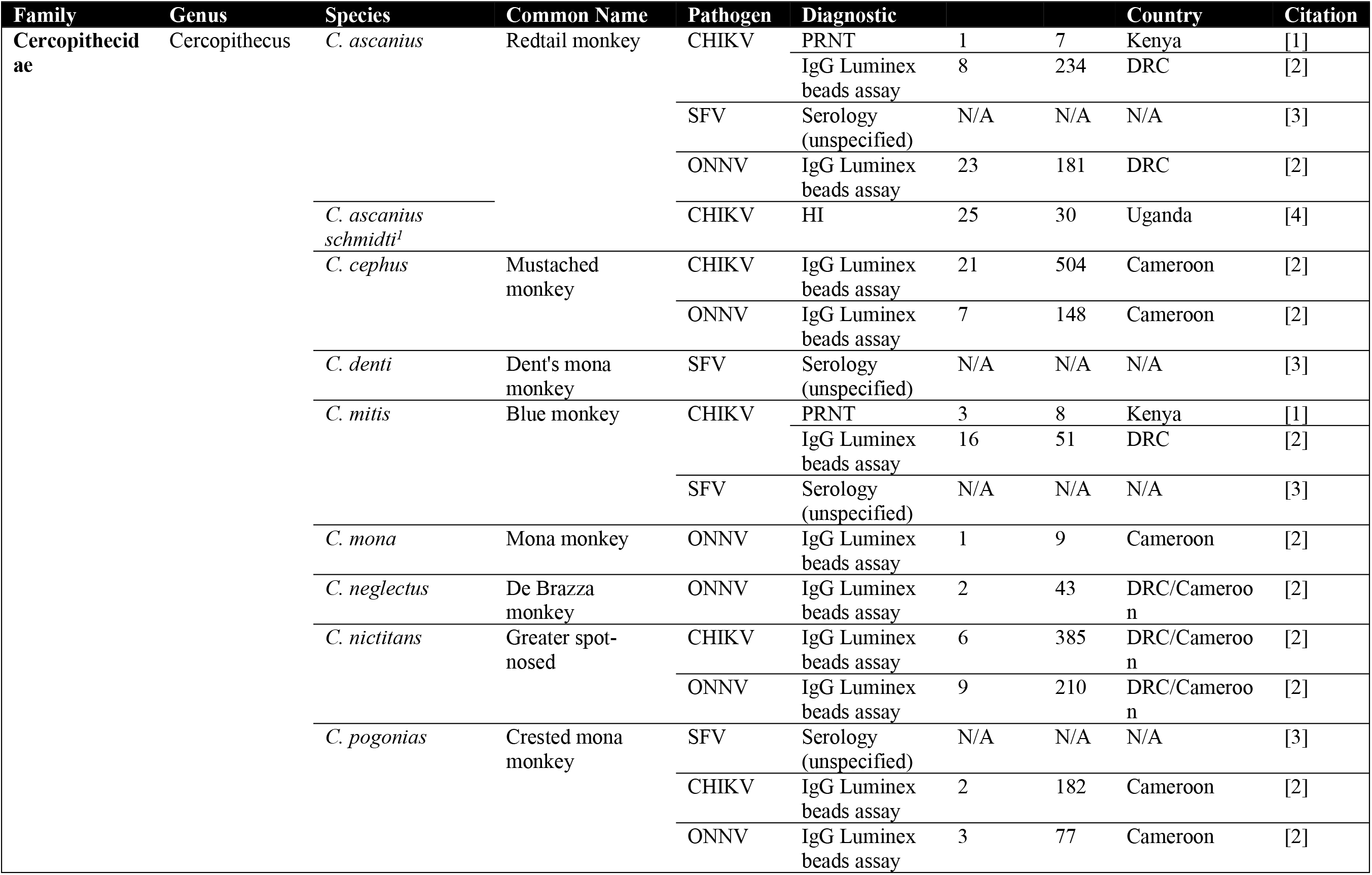
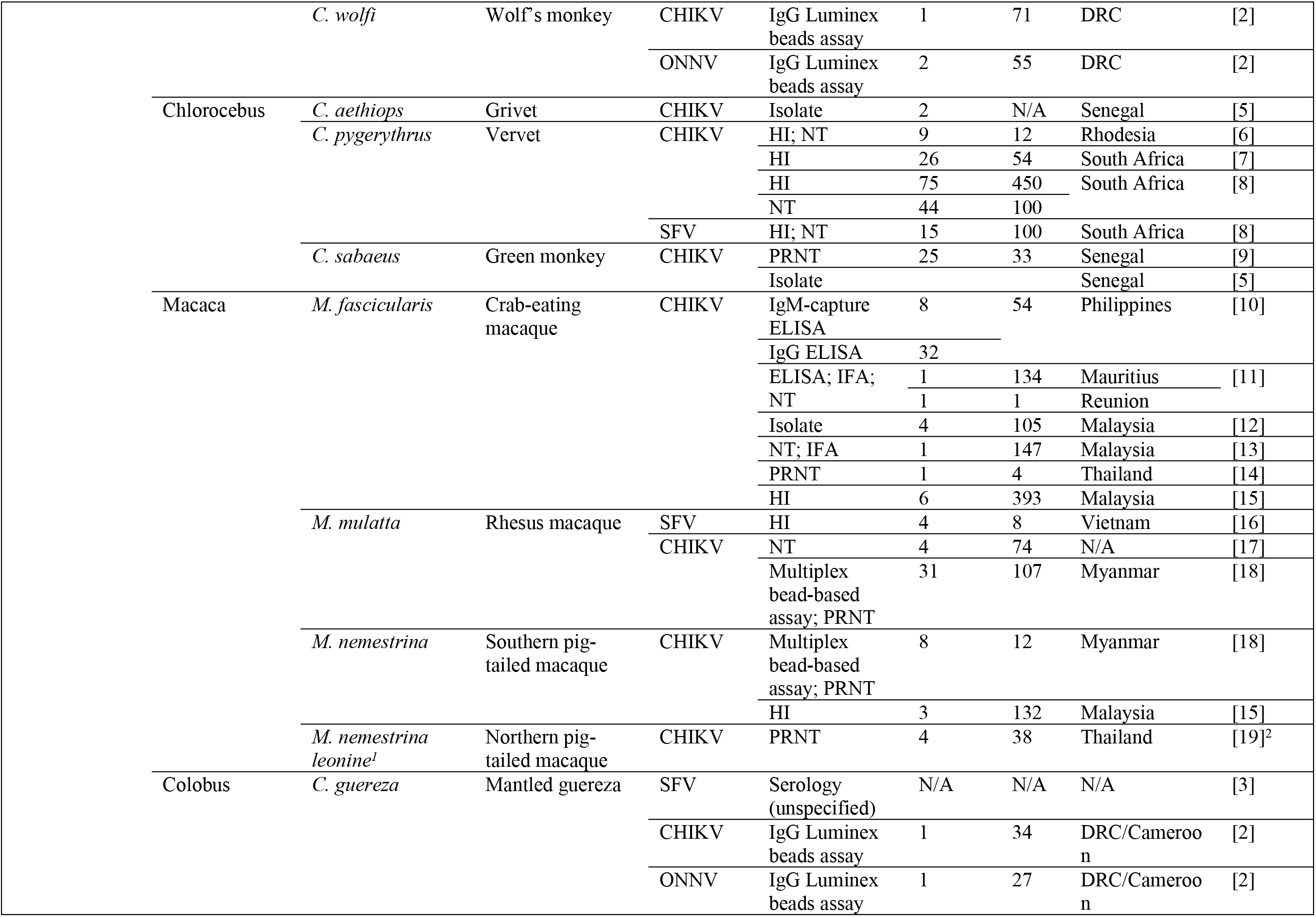
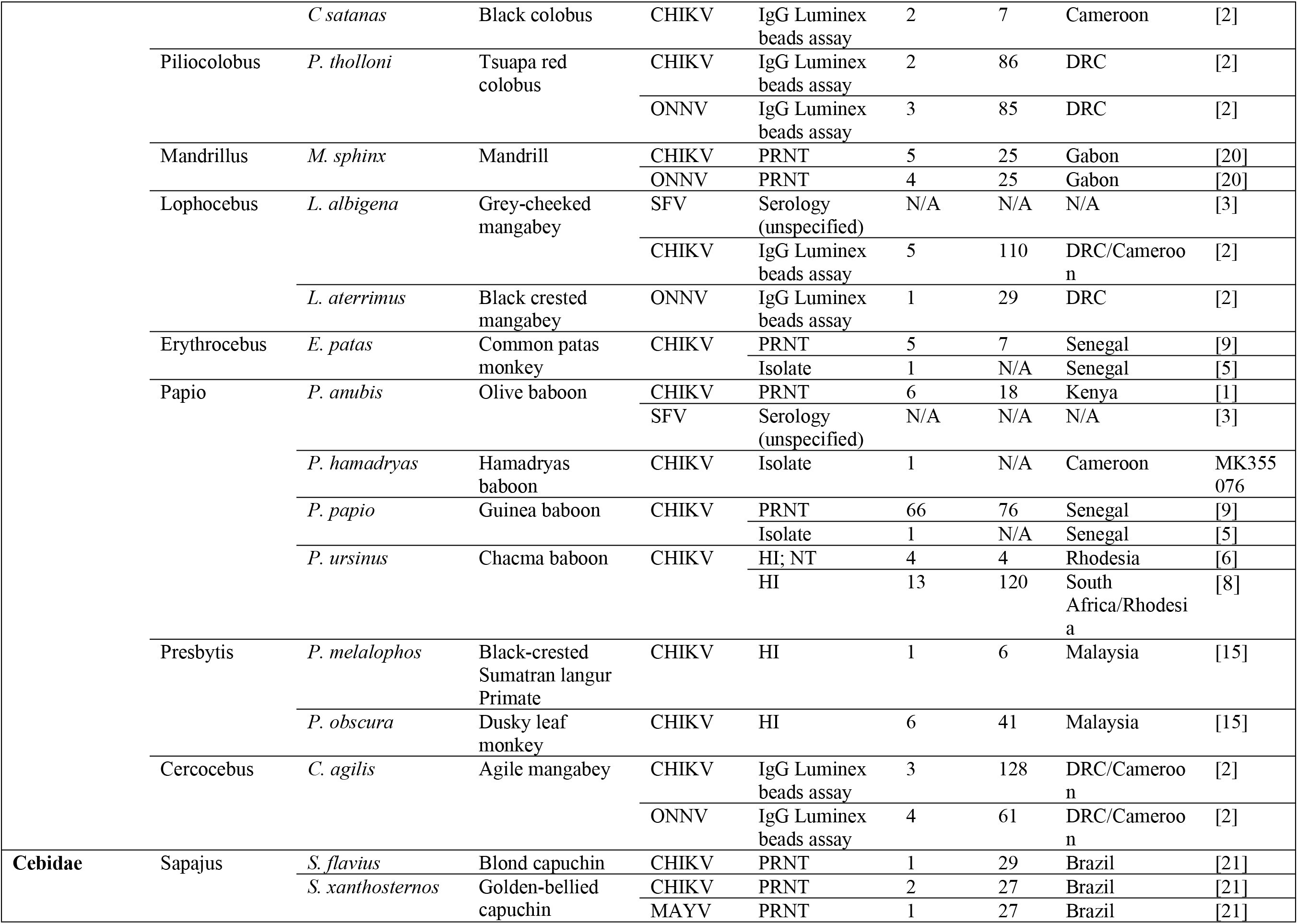
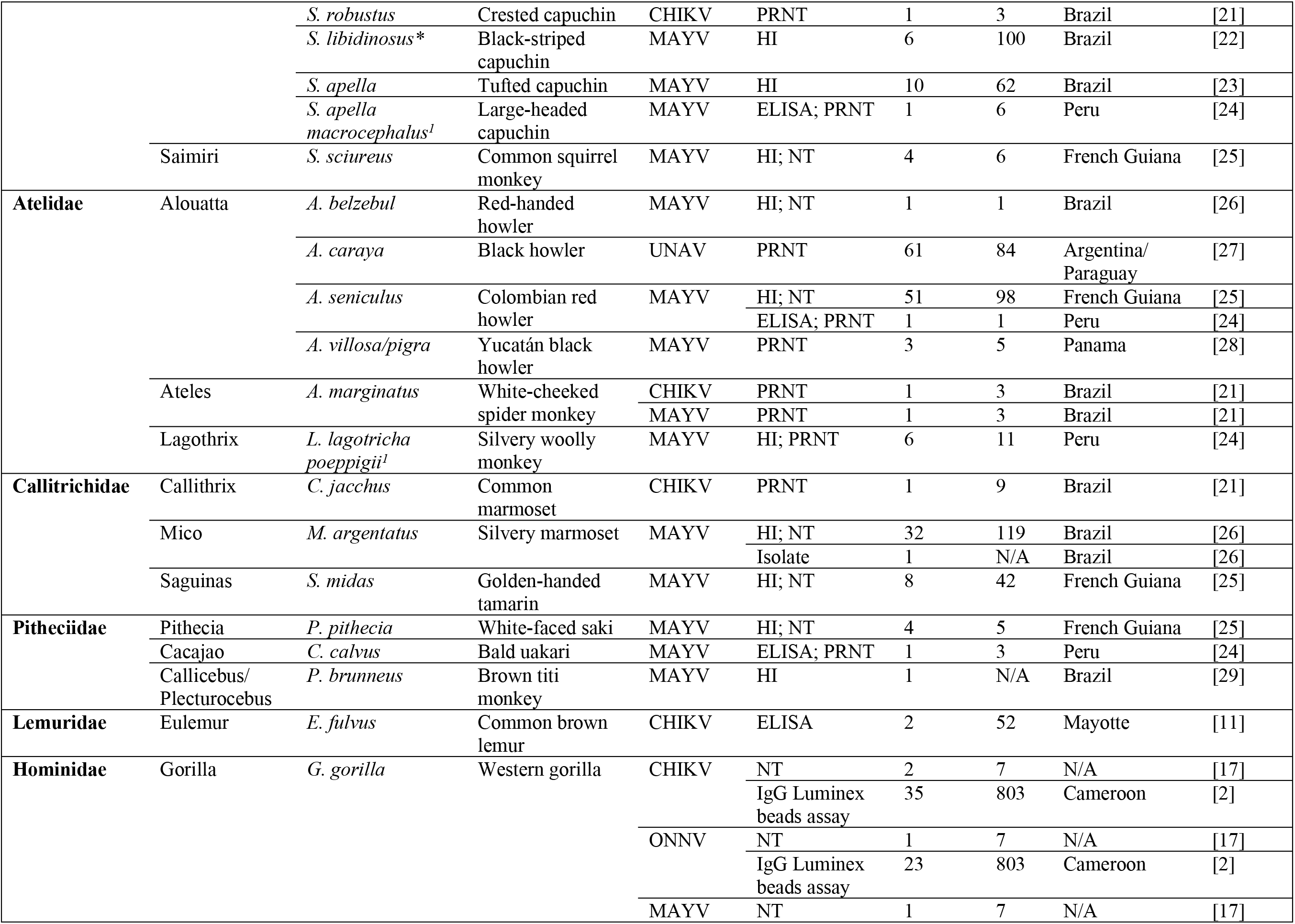
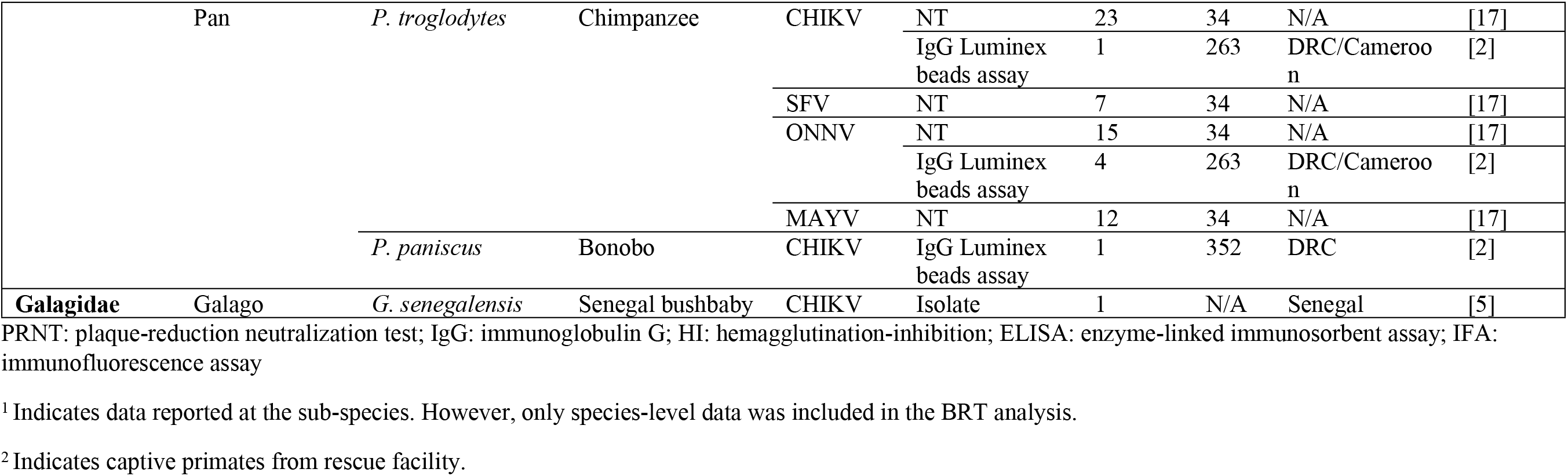
Published records of non-human primate species sampled for *Alphaviruses* using various diagnostic methods.

**Table S2.** Traits considered for BRT model. For each trait, we include the type (taxonomy, morphology, reproduction etc.), a short description, unit, source, and percent coverage. Coverage represents the percentage of total NHP species with available data for a given trait.

| Trait | Type | Description | Units | Source | Coverage (%) |
| --- | --- | --- | --- | --- | --- |
| **species** | Taxonomy | IUCN 2020 binomial | N/A | COMBINE | 100 |
| **adult_mass_g** | Morphology | Body mass of an adult individual | g | COMBINE | 97 |
| **brain_mass_g** | Morphology | Weight of the brain of an adult individual | g | COMBINE | 97 |
| **adult_body_length_mm** | Morphology | Total length from tip of the nose to anus or base of the tail of an adult individual | mm | COMBINE | 97 |
| **max_longevity_d** | Phenology | Maximum reported age at death for the species | days | COMBINE | 97 |
| **maturity_d** | Phenology | The amount of time needed to reach sexual maturity | days | COMBINE | 66 |
| **female_maturity_d** | Phenology | The amount of time needed for a female to reach sexual maturity | days | COMBINE | 97 |
| **male_maturity_d** | Phenology | The amount of time needed for a male to reach sexual maturity | days | COMBINE | 35 |
| **age_first_reproduction_d** | Reproduction | Age at which females give birth to their first litter or their young attach to teats | days | COMBINE | 97 |
| **gestation_length_d** | Reproduction | Length of time of fetal growth | days | COMBINE | 97 |
| **litter_size_n** | Reproduction | Number of offspring born per litter per female | count | COMBINE | 97 |
| **litters_per_year_n** | Reproduction | Number of litters per female per year | count | COMBINE | 97 |
| **interbirth_interval_d** | Reproduction | Time between reproduction events in days | days | COMBINE | 97 |
| **neonate_mass_g** | Morphology | Weight of an individual at birth in grams | g | COMBINE | 60 |
| **weaning_age_d** | Phenology | Age at which primary nutritional dependency on the mother ends and independent foraging begins in days | days | COMBINE | 97 |
| **weaning_mass_g** | Morphology | Weight at weaning in grams | g | COMBINE | 27 |
| **generation_length_d** | Phenology | Average age of parents of the current cohort in days | days | COMBINE | 97 |
| **dispersal_km** | Behavior | The distance an animal travels between its place of birth to the place where it reproduces in kilometers | km | COMBINE | 97 |
| **density_n_km2** | Behavior | Number of individuals of the species per squared kilometer | km2 | COMBINE | 70 |
| **home_range_km2** | Behavior | Size of the area within which everyday activities of individuals or groups of individuals are typically restricted in km2 | km2 | COMBINE | 43 |
| **social_group_n** | Behavior | Number of individuals in a group that spends most of their daily time together | count | COMBINE | 59 |
| **det_inv** | Diet | Percentage of the diet composed of invertebrates | % | COMBINE | 79 |
| **det_vend** | Diet | Percentage of the diet composed of mammals, birds | % | COMBINE | 79 |
| **det_vect** | Diet | Percentage of the diet composed of reptiles, snakes, amphibians, salamanders | % | COMBINE | 79 |
| **det_vunk** | Diet | Percentage of the diet composed of vertebrates – general or unknown | % | COMBINE | 79 |
| **det_fruit** | Diet | Percentage of the diet composed of fruit, drupes | % | COMBINE | 79 |
| **det_nect** | Diet | Percentage of the diet composed of nectar, pollen, plant exudates, gums | % | COMBINE | 79 |
| **det_seed** | Diet | Percentage of the diet composed of seed, maize, nuts, spores, wheat, grains | % | COMBINE | 79 |
| **det_plantother** | Diet | Percentage of the diet composed of other plant elements | % | COMBINE | 79 |
| **det_diet_breadth_n** | Diet | Number of prevalent ($\geq 20\%$) EltonTraits dietary categories consumed | count | COMBINE | 97 |
| **folivore** | Diet | Diet composed of $>60\%$ leaves | binary | ETWP | 90 |
| **folivore_frugivore** | Diet | Diet comprised of both fruits/seeds and leaves in similar proportions | binary | ETWP | 90 |
| **frugivore** | Diet | Diet composed of $>60\%$ of fruits/seeds in diet | binary | ETWP | 90 |
| **insectivore** | Diet | Diet composed of $>50\%$ arthropods | binary | ETWP | 90 |
| **omnivore** | Diet | Diet comprised of both plants and animals in similar proportions | binary | ETWP | 90 |
| **gummivore** | Diet | Diet dominated by plant exudates | binary | ETWP | 90 |
| **foraging_stratum_arboreal** | Behavior | Arboreal foraging | binary | COMBINE | 100 |
| **foraging_stratum_ground** | Behavior | Ground-level foraging | binary | COMBINE | 100 |
| **foraging_stratum_scansorial** | Behavior | Scansorial foraging | binary | COMBINE | 100 |
| **activity_cycle_nocturnal** | Behavior | Nocturnal activity cycle | binary | COMBINE | 100 |
| **activity_cycle_diurnal** | Behavior | Diurnal activity cycle | binary | COMBINE | 100 |
| **activity_cycle_multiple** | Behavior | Activity cycle is nocturnal/crepuscular, cathemeral, crepuscular or diurnal/crepuscular | binary | COMBINE | 100 |
| **upper_elevation_m** | Geography | Upper elevation limit at which the species can be found in meters | m | COMBINE | 40 |
| **lower_elevation_m** | Geography | Lower elevation limit at which the species can be found in meters | m | COMBINE | 29 |
| **altitude_breadth_m** | Geography | Difference between the upper and lower elevation limits of a species in meters | m | COMBINE | 28 |
| **island_dwelling** | Geography | 20% or more of the breeding range occurs on an island | binary | COMBINE | 38 |
| **disected_by_mountains** | Geography | Range dissected by mountains (based on elevation gradients with slopes equal or higher than 5 degrees) | binary | COMBINE | 38 |
| **afrotropical** | Geography | Species can be encountered in Afrotropical biogeographical realm | binary | COMBINE | 100 |
| **australasian** | Geography | Species can be encountered in Australasian biogeographical realm | binary | COMBINE | 100 |
| **indomalayan** | Geography | Species can be encountered in Indomalayan biogeographical realm | binary | COMBINE | 100 |
| **neotropical** | Geography | Species can be encountered in Neotropical biogeographical realm | binary | COMBINE | 100 |
| **paleartic** | Geography | Species can be encountered in Palearctic biogeographical realm | binary | COMBINE | 100 |
| **realm_multiple** | Geography | Species can be encountered in multiple biogeographical realms | binary | COMBINE | 100 |
| **habitat_breadth_n** | Habitat | Number of distinct suitable level 1 IUCN habitats | count | COMBINE | 93 |
| **male_mass_kg** | Morphology | Body mass of an adult male individual | kg | ETWP | 69 |
| **female_mass_kg** | Morphology | Body mass of an adult female individual | kg | ETWP | 67 |
| **habitat_forest** | Habitat | Species resides in IUCN forest habitat type; includes ecosystems such as tropical wet forest, cloud forest, dry forest, montane forest, temperate forest and semideciduous forest | binary | ETWP | 93 |
| **habitat_savannah** | Habitat | Species resides in IUCN savannah habitat type; includes savannah forest and savannah mosaics | binary | ETWP | 93 |
| **habitat_shrubland** | Habitat | Species resides in IUCN shrubland habitat type; includes ecosystems dominated by shrubs, such as scrub, brush and bush | binary | ETWP | 93 |
| **habitat_grassland** | Habitat | Species resides in IUCN grassland habitat type; includes ecosystems mainly composed of grasses and other herbaceous plants; | binary | ETWP | 93 |
| **habitat_wetlands** | Habitat | Species resides in IUCN wetlands habitat type; includes ecosystems such as swamps, flooded forest, swampy forest, wetlands and mangroves | binary | ETWP | 93 |
| **habitats_rocky_areas** | Habitat | Species resides in IUCN rocky areas habitat type; includes ecosystems such as inland cliffs and mountain peaks | binary | ETWP | 93 |
| **habitat_desert** | Habitat | Species resides in IUCN desert habitat type | binary | ETWP | 93 |
| **threat_critical** | Conservation | Species is in the critically endangered IUCN category | binary | ETWP | 93 |
| **threat_endangered** | Conservation | Species is in the endangered IUCN category | binary | ETWP | 93 |
| **threat_vulnerable** | Conservation | Species is in the vulnerable IUCN category | binary | ETWP | 93 |
| **threat_near_threatened** | Conservation | Species is in the near threatened IUCN category | binary | ETWP | 93 |
| **threat_least_concern** | Conservation | Species is in the least concern IUCN category | binary | ETWP | 93 |
| **threat_data_deficient** | Conservation | Species is in the data deficient IUCN category | binary | ETWP | 93 |
| **population_increasing** | Conservation | Population trend increasing according to IUCN | binary | ETWP | 93 |
| **population_stable** | Conservation | Population trend stable according to IUCN | binary | ETWP | 93 |
| **population_decreasing** | Conservation | Population trend decreasing according to IUCN | binary | ETWP | 93 |
| **locomotion_terrestrial** | Behavior | Primate species are frequently active on the ground as well as in trees | binary | ETWP | 89 |
| **locomotion_arboreal** | Behavior | Strictly arboreal locomotion type whereby primate species rarely go to the ground | binary | ETWP | 89 |
| **locomotion_both** | Behavior | Terrestrial locomotion type whereby primate species carry out most of their daily activity on the ground | binary | ETWP | 89 |
| **max_latitude_WGS84dd** | Geography | Maximum latitude of the species' range | decimal degrees | IUCN RedList | 98 |
| **min_latitude_WGS84dd** | Geography | Minimum latitude of the species' range | decimal degrees | IUCN RedList | 98 |
| **range_latitude_WGS84dd** | Geography | Span of latitude covered by the species' range | decimal degrees | IUCN RedList | 98 |
| **centroid_latitude_WGS84dd** | Geography | Latitude of the centroid of the species' range | decimal degrees | IUCN RedList | 98 |
| **mammal_richness_mean** | Habitat | Average mammal species richness within the NHP species' range | count | SEDAC | 98 |
| **human_footprint_mean** | Human | Mean human footprint value within the species' range | continuous (low – high) | SEDAC | 98 |
| **mass_specific_production** | Derived | Mean mass of offspring produced per year normalized by adult body size |  | This study | 60 |
| **postnatal_growth_rate_g** | Derived | Weaning body mass/neonatal body mass | g | This study | 26 |
| **home_range_scaled_km2_g** | Derived | Home range size scaled by adult body mass | km<sup>2</sup>/g | This study | 43 |
| **rel_sexual_maturity_days** | Behavior | Relative sexual maturity age [i.e., age at sexual maturity/maximum longevity (days)] | days | This study | 66 |
| **rel_age_fb_days** | Behavior | Relative age at first birth [age at first birth/maximum longevity (days)] | days | This study | 97 |
| **body_size_ratio_g** |  | Adult Body Mass (g)/Neonate Body Mass (g) | g | This study | 60 |
| **temp_mean_dC** | Climate | Mean temperature documented within the species' range | Celsius | WorldClim | 98 |
| **temp_min_dC** | Climate | Minimum temperature within a species' range | Celsius | WorldClim | 98 |
| **temp_max_dC** | Climate | Maximum temperature within a species' range | Celsius | WorldClim | 98 |
| **temp_range_dC** | Climate | Temperature range within a species' range | Celsius | WorldClim | 98 |
| **precip_mean_mm** | Climate | Mean precipitation within a species' range | mm | WorldClim | 98 |
| **precip_min_mm** | Climate | Minimum precipitation within a species' range | mm | WorldClim | 98 |
| **precip_max_mm** | Climate | Maximum precipitation within a species' range | mm | WorldClim | 98 |
| **precip_range_mm** | Climate | Precipitation range within a species' range | mm | WorldClim | 98 |
| **human_pop_dens_mean** | Human | Mean human population density within a species' range | mean | SEDAC | 98 |
| **human_pop_dens_min** | Human | Minimum human population density in the species' range | Min | SEDAC | 98 |
| **human_pop_dens_max** | Human | Maximum human population density in the species' range | Max | SEDAC | 98 |
| **human_pop_dens_median** | Human | Median human population density in the species' range | median | SEDAC | 98 |
| **ecoregions_n** | Habitat | Number of distinct ecoregions within the species range | Count |  | 98 |
| **cheirogaleidae** | Taxonomy | Species belongs to cheirogaleidae family | binary | IUCN RedList | 100 |
| **galagidae** | Taxonomy | Species belongs to galagidae family | binary | IUCN RedList | 100 |
| **tarsiidae** | Taxonomy | Species belongs to tarsiidae family | binary | IUCN RedList | 100 |
| **callitrichidae** | Taxonomy | Species belongs to callitrichidae family | binary | IUCN RedList | 100 |
| **lorisidae** | Taxonomy | Species belongs to lorisidae family | binary | IUCN RedList | 100 |
| **lepilemuridae** | Taxonomy | Species belongs to lepilemuridae family | binary | IUCN RedList | 100 |
| **indriidae** | Taxonomy | Species belongs to indriidae family | binary | IUCN RedList | 100 |
| **lemuridae** | Taxonomy | Species belongs to lemuridae family | binary | IUCN RedList | 100 |
| **aotidae** | Taxonomy | Species belongs to aotidae family | binary | IUCN RedList | 100 |
| **pitheciidae** | Taxonomy | Species belongs to pitheciidae family | binary | IUCN RedList | 100 |
| **atelidae** | Taxonomy | Species belongs to atelidae family | binary | IUCN RedList | 100 |
| **cebidae** | Taxonomy | Species belongs to cebidae family | binary | IUCN RedList | 100 |
| **cercopithecidae** | Taxonomy | Species belongs to cercopithecidae family | binary | IUCN RedList | 100 |
| **daubentoniidae** | Taxonomy | Species belongs to daubentoniidae family | binary | IUCN RedList | 100 |
| **hylobatidae** | Taxonomy | Species belongs to hylobatidae family | binary | IUCN RedList | 100 |
| **palaeopropithecidae** | Taxonomy | Species belongs to palaeopropithecidae family | binary | IUCN RedList | 100 |
| **hominidae** | Taxonomy | Species belongs to hominidae family | binary | IUCN RedList | 100 |

ETWP: Ecological Traits of the World's Primates; COMBINE: COalesced Mammal dataBase of INtrinsic and Extrinsic traits; IUCN: International Union for the Conservation of Nature; SEDAC: Socioeconomic Data and Applications Center

**Table S3.**
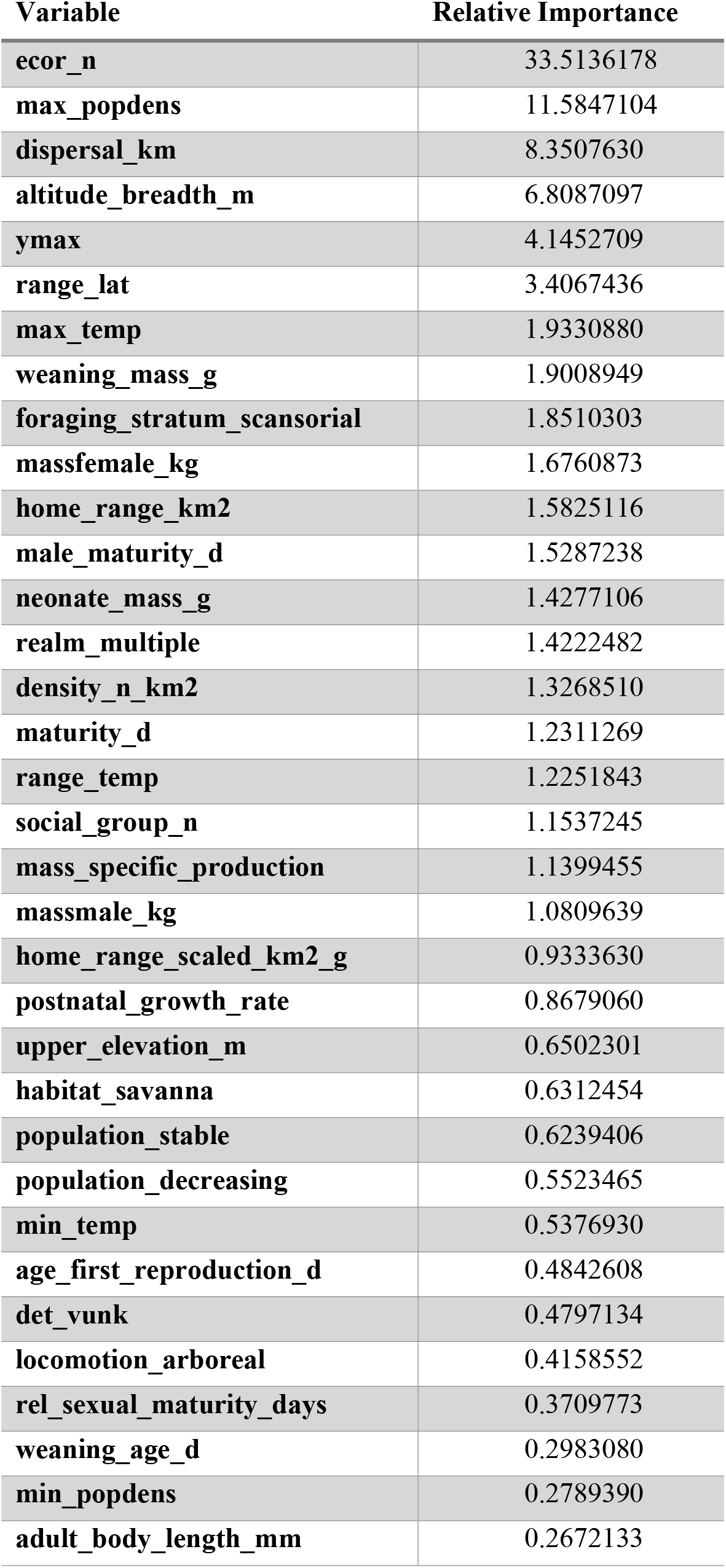
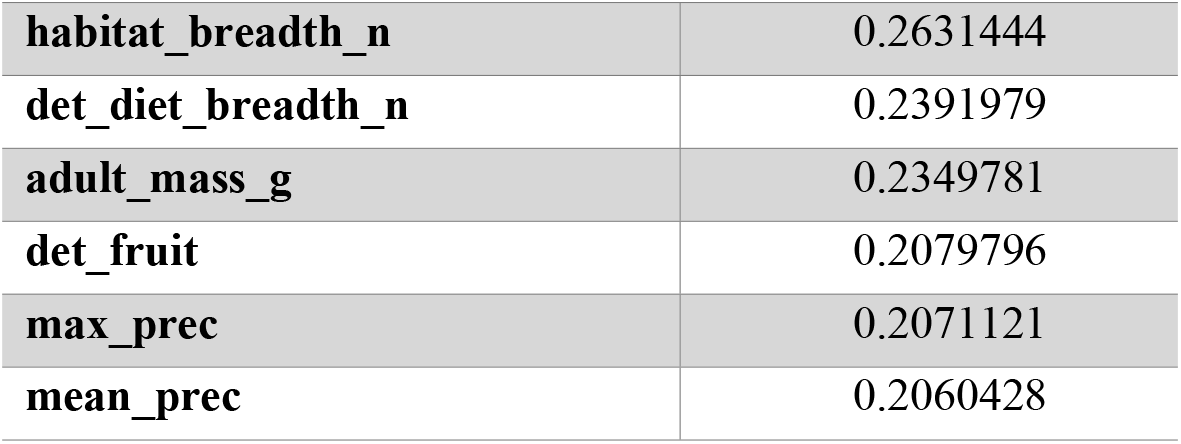
Relative importance scores for variables predicting study effort. Relative importance scores for variables included in the boosted regression tree model, with Web of Science citation count used as the outcome variable. The 40 variables with the highest relative importance are included in the table.

**Table S4.** Relative importance scores for variables included in main analysis. Relative importance scores from the boosted regression tree model predicting *Alphavirus* host status.

| Variable | Relative importance |
| --- | --- |
| range_lat | 13.889279 |
| max_longevity_d | 5.950387 |
| max_temp | 5.262230 |
| min_popdens | 5.089114 |
| ecor_n | 4.858153 |
| neonate_mass_g | 4.262735 |
| massfemale_kg | 4.224301 |
| mean_prec | 4.142488 |
| home_range_km2 | 3.973843 |
| social_group_n | 3.636380 |
| hfp_avg | 3.314835 |
| weaning_age_d | 3.201236 |
| ymin | 3.099195 |
| min_temp | 2.878069 |
| massmale_kg | 2.871080 |
| rel_sexual_maturity_days | 2.633169 |
| det_fruit | 2.608860 |
| home_range_scaled_km2_g | 2.589837 |
| density_n_km2 | 2.527599 |
| age_first_reproduction_d | 2.166246 |
| maturity_d | 2.007293 |
| body_size_ratio | 1.965063 |
| max_popdens | 1.898989 |
| generation_length_d | 1.890021 |
| det_vunk | 1.831010 |
| mean_popdens | 1.797108 |
| min_prec | 1.740699 |
| **range_prec** | 1.365326 |
| **island_dwelling** | 1.189970 |
| **postnatal_growth_rate** | 1.135486 |

**Table S5.**
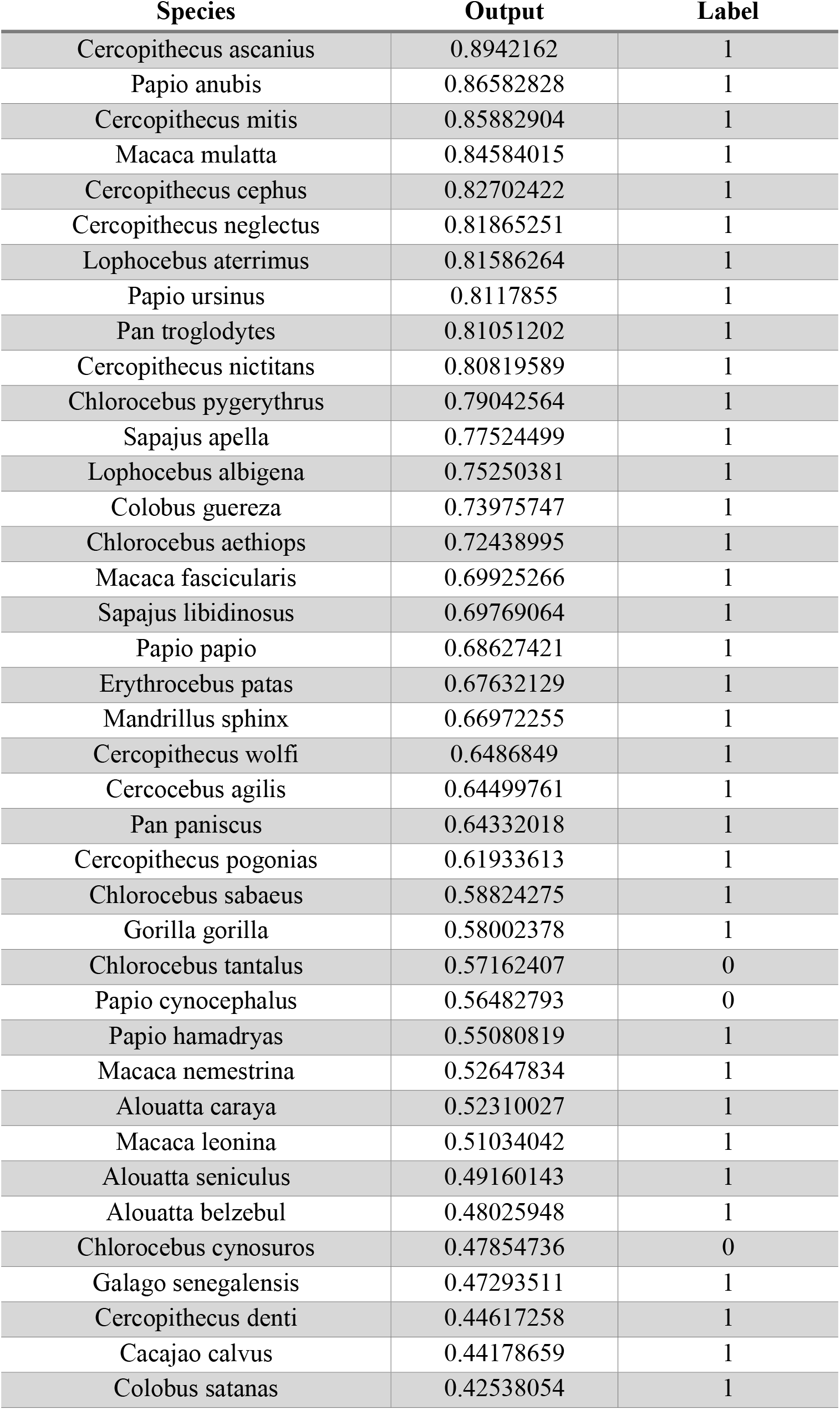
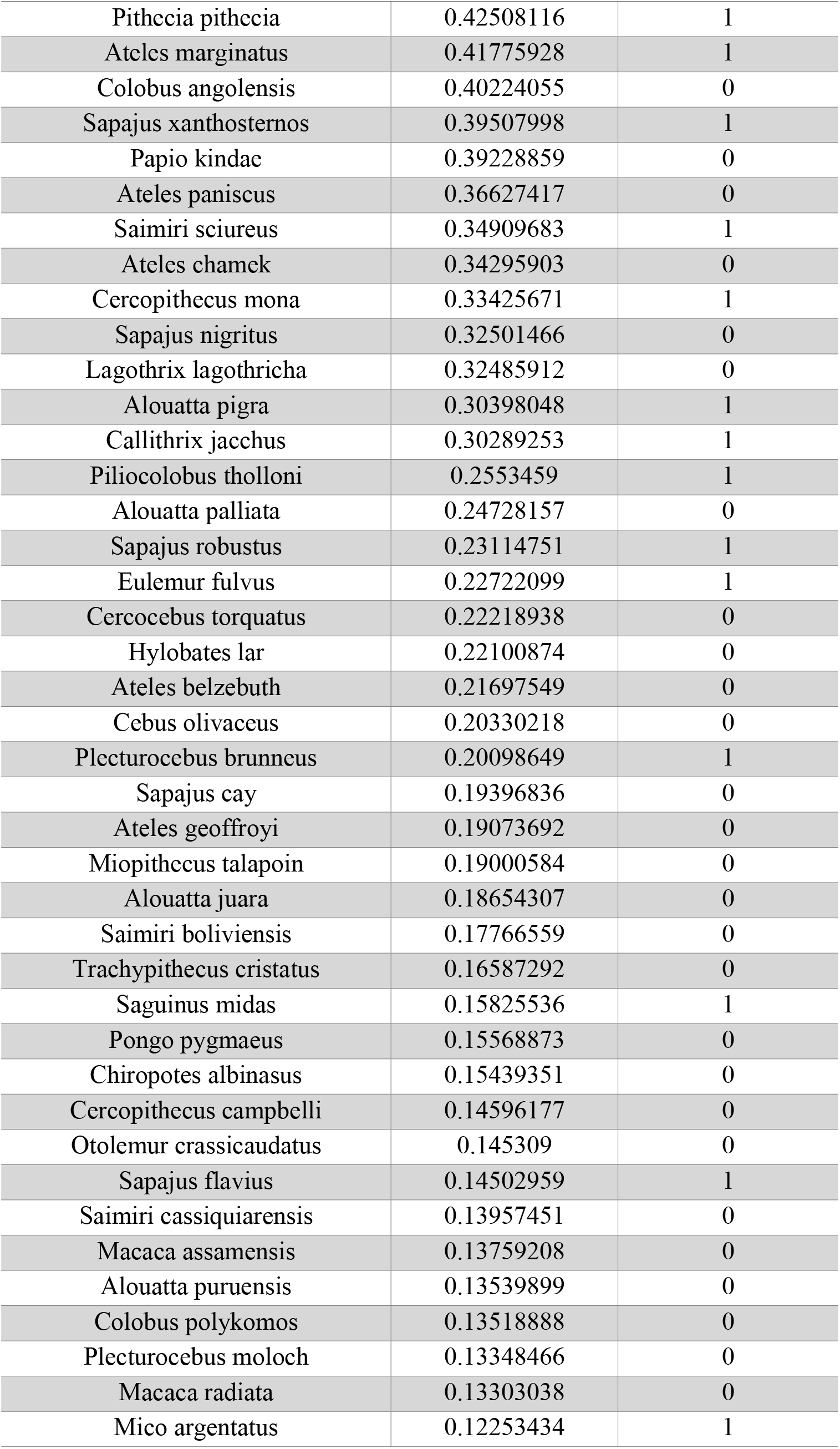
List of NHP species and predicted probability of carrier status. This table contains the 80 non-human primate species with the highest predicted probability of carrier status. The label variable denotes known *Alphavirus* carriers. The corrected AUC of our model was 0.91, indicating good discrimination between known hosts and non-hosts after accounting for the null model performance.

## References

1. Cunha MS, Costa PAG, Correa IA, de Souza MRM, Calil PT, da Silva GPD, et al. Chikungunya Virus: An Emergent Arbovirus to the South American Continent and a Continuous Threat to the World. Frontiers in microbiology. 2020;11:1297. Epub 2020/07/17. doi: 10.3389/fmicb.2020.01297. PubMed PMID: 32670231; PubMed Central PMCID: PMCPMC7332961.

2. LeDuc JW, Pinheiro FP, Travassos da Rosa AP. An outbreak of Mayaro virus disease in Belterra, Brazil. II. Epidemiology. Am J Trop Med Hyg. 1981;30(3):682–8. Epub 1981/05/01. doi: 10.4269/ajtmh.1981.30.682. PubMed PMID: 6266264.

3. Schmaljohn AL, McClain D. Alphaviruses (Togaviridae) and Flaviviruses (Flaviviridae). In: th, Baron S, editors. Medical Microbiology. Galveston (TX): University of Texas Medical Branch at Galveston; 1996.

4. Levi LI, Vignuzzi M. Arthritogenic Alphaviruses: A Worldwide Emerging Threat? Microorganisms. 2019;7(5). Epub 2019/05/17. doi: 10.3390/microorganisms7050133. PubMed PMID: 31091828; PubMed Central PMCID: PMCPMC6560413.

5. Suhrbier A, Jaffar-Bandjee MC, Gasque P. Arthritogenic alphaviruses--an overview. Nat Rev Rheumatol. 2012;8(7):420-9. Epub 2012/05/09. doi: 10.1038/nrrheum.2012.64. PubMed PMID: 22565316.

6. De O’Mota MT, Avilla CMS, Nogueira ML. Mayaro virus: A neglected threat could cause the next worldwide viral epidemic. Future Virology. 2019;14(6):375–7. doi: 10.2217/fvl-2019-0051.

7. Sun JF, Wu D. Mayaro virus, a regional or global threat? Travel Medicine and Infectious Disease. 2019;32. doi: 10.1016/j.tmaid.2019.07.018. PubMed PMID: WOS:000502623800021.

8. Powers AM, Brault AC, Shirako Y, Strauss EG, Kang W, Strauss JH, et al. Evolutionary relationships and systematics of the alphaviruses. Journal of virology. 2001;75(21):10118–31. Epub 2001/10/03. doi: 10.1128/jvi.75.21.10118-10131.2001. PubMed PMID: 11581380; PubMed Central PMCID: PMCPMC114586.

9. Ganjian N, Riviere-Cinnamond A. Mayaro virus in Latin America and the Caribbean. Revista panamericana de salud publica = Pan American journal of public health. 2020;44:e14. Epub 2020/02/14. doi: 10.26633/rpsp.2020.14. PubMed PMID: 32051685; PubMed Central PMCID: PMCPMC7008609.

10. Harley D, Sleigh A, Ritchie S. Ross River virus transmission, infection, and disease: a cross-disciplinary review. Clin Microbiol Rev. 2001;14(4):909–32, table of contents. Epub 2001/10/05. doi: 10.1128/cmr.14.4.909-932.2001. PubMed PMID: 11585790; PubMed Central PMCID: PMCPMC89008.

11. Pezzi L, LaBeaud AD, Reusken CB, Drexler JF, Vasilakis N, Diallo M, et al. GloPID-R report on chikungunya, o’nyong-nyong and Mayaro virus, part 2: Epidemiological distribution of o’nyong-nyong virus. Antiviral Res. 2019;172:104611. Epub 2019/09/24. doi: 10.1016/j.antiviral.2019.104611. PubMed PMID: 31545982.

12. Celone M, Okech B, Han BA, Forshey BM, Anyamba A, Dunford J, et al. A systematic review and meta-analysis of the potential non-human animal reservoirs and arthropod vectors of the Mayaro virus. PLoS Negl Trop Dis. 2021;15(12):e0010016. Epub 2021/12/14. doi: 10.1371/journal.pntd.0010016. PubMed PMID: 34898602; PubMed Central PMCID: PMCPMC8699665.

13. Mongkol N, Wang FS, Suthisawat S, Likhit O, Charoen P, Boonnak K. Seroprevalence of Chikungunya and Zika virus in nonhuman primates: A systematic review and meta-analysis. One Health. 2022;15:100455. Epub 2022/12/20. doi: 10.1016/j.onehlt.2022.100455. PubMed PMID: 36532673; PubMed Central PMCID: PMCPMC9754931.

14. Mackenzie JS, Jeggo M. Reservoirs and vectors of emerging viruses. Current opinion in virology. 2013;3(2):170–9.

15. Bueno MG, Martinez N, Abdalla L, Duarte Dos Santos CN, Chame M. Animals in the Zika Virus Life Cycle: What to Expect from Megadiverse Latin American Countries. PLoS Negl Trop Dis. 2016;10(12):e0005073. Epub 2016/12/23. doi: 10.1371/journal.pntd.0005073. PubMed PMID: 28005902; PubMed Central PMCID: PMCPMC5179043.

16. Komar N. West Nile virus surveillance using sentinel birds. Ann N Y Acad Sci. 2001;951:58–73. Epub 2002/01/19. doi: 10.1111/j.1749-6632.2001.tb02685.x. PubMed PMID: 11797805.

17. Cunha MS, da Costa AC, de Azevedo Fernandes NCC, Guerra JM, Dos Santos FCP, Nogueira JS, et al. Epizootics due to yellow fever virus in São Paulo State, Brazil: viral dissemination to new areas (2016– 2017). Scientific reports. 2019;9(1):1-13.

18. de Oliveira Figueiredo P, Stoffella-Dutra AG, Barbosa Costa G, Silva de Oliveira J, Dourado Amaral C, Duarte Santos J, et al. Re-Emergence of Yellow Fever in Brazil during 2016-2019: Challenges, Lessons Learned, and Perspectives. Viruses. 2020;12(11). Epub 2020/11/05. doi: 10.3390/v12111233. PubMed PMID: 33143114; PubMed Central PMCID: PMCPMC7692154.

19. Yang XL, Zhang YZ, Jiang RD, Guo H, Zhang W, Li B, et al. Genetically Diverse Filoviruses in Rousettus and Eonycteris spp. Bats, China, 2009 and 2015. Emerg Infect Dis. 2017;23(3):482-6. Epub 2017/02/22. doi: 10.3201/eid2303.161119. PubMed PMID: 28221123; PubMed Central PMCID: PMCPMC5382765.

20. Carlson CJ, Gibb RJ, Albery GF, Brierley L, Connor RP, Dallas TA, et al. The Global Virome in One Network (VIRION): an Atlas of Vertebrate-Virus Associations. mBio. 2022;13(2):e0298521. Epub 2022/03/02. doi: 10.1128/mbio.02985-21. PubMed PMID: 35229639; PubMed Central PMCID: PMCPMC8941870.

21. Haydon DT, Cleaveland S, Taylor LH, Laurenson MK. Identifying reservoirs of infection: a conceptual and practical challenge. Emerg Infect Dis. 2002;8(12):1468–73. Epub 2002/12/25. doi: 10.3201/eid0812.010317. PubMed PMID: 12498665; PubMed Central PMCID: PMCPMC2738515.

22. Berger SA. GIDEON: a comprehensive Web-based resource for geographic medicine. Int J Health Geogr. 2005;4(1):10. Epub 2005/04/26. doi: 10.1186/1476-072x-4-10. PubMed PMID: 15847698; PubMed Central PMCID: PMCPMC1090610.

23. Han BA, Schmidt JP, Bowden SE, Drake JM. Rodent reservoirs of future zoonotic diseases. Proceedings of the National Academy of Sciences of the United States of America. 2015;112(22):7039–44. Epub 2015/06/04. doi: 10.1073/pnas.1501598112. PubMed PMID: 26038558; PubMed Central PMCID: PMCPMC4460448.

24. Han BA, Schmidt JP, Alexander LW, Bowden SE, Hayman DT, Drake JM. Undiscovered Bat Hosts of Filoviruses. PLoS Negl Trop Dis. 2016;10(7):e0004815. Epub 2016/07/16. doi: 10.1371/journal.pntd.0004815. PubMed PMID: 27414412; PubMed Central PMCID: PMCPMC4945033.

25. Han BA, Majumdar S, Calmon FP, Glicksberg BS, Horesh R, Kumar A, et al. Confronting data sparsity to identify potential sources of Zika virus spillover infection among primates. Epidemics. 2019;27:59–65. Epub 2019/03/25. doi: 10.1016/j.epidem.2019.01.005. PubMed PMID: 30902616.

26. Soria CD, Pacifici M, Di Marco M, Stephen SM, Rondinini C. COMBINE: a coalesced mammal database of intrinsic and extrinsic traits. Ecology. 2021;102(6):e03344. Epub 2021/03/21. doi: 10.1002/ecy.3344. PubMed PMID: 33742448.

27. Galán-Acedo C, Arroyo-Rodríguez V, Andresen E, Arasa-Gisbert R. Ecological traits of the world’s primates. Sci Data. 2019;6(1):55. Epub 2019/05/16. doi: 10.1038/s41597-019-0059-9. PubMed PMID: 31086194; PubMed Central PMCID: PMCPMC6513815.

28. Lumbierres M, Dahal PR, Soria CD, Di Marco M, Butchart SHM, Donald PF, et al. Area of Habitat maps for the world’s terrestrial birds and mammals. Scientific Data. 2022;9(1):749. doi: 10.1038/s41597-022-01838-w.

29. Fick SE, Hijmans RJ. WorldClim 2: new 1km spatial resolution climate surfaces for global land areas. International Journal of Climatology. 2017;37(12):4302–15.

30. Center for International Earth Science Information Network - CIESIN - Columbia University. Gridded Population of the World, Version 4 (GPWv4): Population Density, Revision 11. Palisades, New York: NASA Socioeconomic Data and Applications Center (SEDAC); 2018.

31. IUCN (International Union for the Conservation of Nature). In: 2024(1) RLSRRsdTIRLoTS, editor. 2024-1 ed. https://www.iucnredlist.org Accessed on June 20, 2024.

32. Sanderson EW, Jaiteh M, Levy MA, Redford KH, Wannebo AV, Woolmer G. The Human Footprint and The Last of the Wild. BioScience. 2002;52(10):891–904.

33. Olson DM, Dinerstein E, Wikramanayake ED, Burgess ND, Powell GV, Underwood EC, et al. Terrestrial Ecoregions of the World: A New Map of Life on Earth: A new global map of terrestrial ecoregions provides an innovative tool for conserving biodiversity. BioScience. 2001;51(11):933–8.

34. IUCN (nternational Union for Conservation of Nature) 2017. IUCN Terrestrial Mammals. The IUCN Red List of Threatened Species. http://www.iucnredlist.org/technical-documents/spatial-data. Downloaded on 01 January 2020. 6.2 ed.

35. Hamilton MJ, Davidson AD, Sibly RM, Brown JH. Universal scaling of production rates across mammalian lineages. Proceedings of the Royal Society B: Biological Sciences. 2011;278(1705):560–6.

36. Plowright RK, Becker DJ, Crowley DE, Washburne AD, Huang T, Nameer PO, et al. Prioritizing surveillance of Nipah virus in India. PLoS Negl Trop Dis. 2019;13(6):e0007393. Epub 2019/06/28. doi: 10.1371/journal.pntd.0007393. PubMed PMID: 31246966; PubMed Central PMCID: PMCPMC6597033.

37. Pandit PS, Doyle MM, Smart KM, Young CCW, Drape GW, Johnson CK. Predicting wildlife reservoirs and global vulnerability to zoonotic Flaviviruses. Nat Commun. 2018;9(1):5425. Epub 2018/12/24. doi: 10.1038/s41467-018-07896-2. PubMed PMID: 30575757; PubMed Central PMCID: PMCPMC6303316.

38. Yang LH, Han BA. Data-driven predictions and novel hypotheses about zoonotic tick vectors from the genus Ixodes. BMC Ecol. 2018;18(1):7. Epub 2018/02/17. doi: 10.1186/s12898-018-0163-2. PubMed PMID: 29448923; PubMed Central PMCID: PMCPMC5815220.

39. Evans MV, Dallas TA, Han BA, Murdock CC, Drake JM. Data-driven identification of potential Zika virus vectors. Elife. 2017;6. Epub 2017/03/01. doi: 10.7554/eLife.22053. PubMed PMID: 28244371; PubMed Central PMCID: PMCPMC5342824.

40. Martin JT, Fischhoff IR, Castellanos AA, Han BA. Ecological Predictors of Zoonotic Vector Status Among Dermacentor Ticks (Acari: Ixodidae): A Trait-Based Approach. J Med Entomol. 2022;59(6):2158–66. Epub 2022/09/07. doi: 10.1093/jme/tjac125. PubMed PMID: 36066562; PubMed Central PMCID: PMCPMC9667724.

41. Vadmal GM, Glidden CK, Han BA, Carvalho BM, Castellanos AA, Mordecai EA. Data-driven predictions of potential Leishmania vectors in the Americas. PLoS Negl Trop Dis. 2023;17(2):e0010749. Epub 2023/02/23. doi: 10.1371/journal.pntd.0010749. PubMed PMID: 36809249; PubMed Central PMCID: PMCPMC9983874.

42. Becker DJ, Albery GF, Sjodin AR, Poisot T, Bergner LM, Chen B, et al. Optimising predictive models to prioritise viral discovery in zoonotic reservoirs. Lancet Microbe. 2022;3(8):e625–e37. Epub 2022/01/18. doi: 10.1016/s2666-5247(21)00245-7. PubMed PMID: 35036970; PubMed Central PMCID: PMCPMC8747432.

43. Fischhoff IR, Castellanos AA, Rodrigues J, Varsani A, Han BA. Predicting the zoonotic capacity of mammals to transmit SARS-CoV-2. Proc Biol Sci. 2021;288(1963):20211651. Epub 2021/11/18. doi: 10.1098/rspb.2021.1651. PubMed PMID: 34784766; PubMed Central PMCID: PMCPMC8596006.

44. Elith J, Leathwick, J.R., Hastie, T. A working guide to boosted regression trees. J Anim Ecol. 2008;77:802–13. doi: 10.1111/j.1365-2656.2008.01390.x.

45. Greenwell B, Boehmke B, Cunningham J. 2019 gbm: Generalized Boosted Regression Models. R package version 2.1.5.

46. Elder J. Evaluate the Validity of Your Discovery with Target Shuffling. White Paper. 2014. Available: https://www.elderresearch.com/wp-content/uploads/2021/01/White-Paper_Evaluate-the-Validity-of-Your-Discovery-with-Target-Shuffling_2021.pdf.

47. Ryser-Degiorgis MP. Wildlife health investigations: needs, challenges and recommendations. BMC Vet Res. 2013;9:223. Epub 2013/11/06. doi: 10.1186/1746-6148-9-223. PubMed PMID: 24188616; PubMed Central PMCID: PMCPMC4228302.

48. Goldstein T, Anthony SJ, Gbakima A, Bird BH, Bangura J, Tremeau-Bravard A, et al. The discovery of Bombali virus adds further support for bats as hosts of ebolaviruses. Nat Microbiol. 2018;3(10):1084–9. Epub 2018/08/29. doi: 10.1038/s41564-018-0227-2. PubMed PMID: 30150734; PubMed Central PMCID: PMCPMC6557442.

49. IUCN. 2026 In: 2026-1. TIRLoTSV, editor. https://www.iucnredlist.org. Accessed on 31 March 2026.

50. Hahn NE, Proulx D, Muruthi PM, Alberts S, Altmann J. Gastrointestinal Parasites in Free-ranging Kenyan Baboons (*Papio cynocephalus* and *P. anubis*). International Journal of Primatology. 2003;24:271–9.

51. Bloomfield LSP, McIntosh TL, Lambin EF. Habitat fragmentation, livelihood behaviors, and contact between people and nonhuman primates in Africa. Landscape Ecology. 2020;35(4):985–1000. doi: 10.1007/s10980-020-00995-w.

52. Nunes VF, Lopes PM, Ferreira RG. Monkeying around Anthropocene: Patterns of human-nonhuman primates’ interactions in Brazil. Ethnobiology and Conservation. 2021;10.

53. Fischer C, Jo WK, Haage V, Moreira-Soto A, de Oliveira Filho EF, Drexler JF. Challenges towards serologic diagnostics of emerging arboviruses. Clin Microbiol Infect. 2021;27(9):1221–9. Epub 2021/06/11. doi: 10.1016/j.cmi.2021.05.047. PubMed PMID: 34111589.

54. Kasbergen LMR, Nieuwenhuijse DF, de Bruin E, Sikkema RS, Koopmans MPG. The increasing complexity of arbovirus serology: An in-depth systematic review on cross-reactivity. PLoS Negl Trop Dis. 2023;17(9):e0011651. Epub 2023/09/22. doi: 10.1371/journal.pntd.0011651. PubMed PMID: 37738270; PubMed Central PMCID: PMCPMC10550177.

55. Weller AK, Chapman OS, Gora SL, Guralnick RP, McLean BS. New insight into drivers of mammalian litter size from individual-level traits. Ecography. 2024;2024(1):e06928. doi: 10.1111/ecog.06928.

56. Kuno G, Chang GJ. Biological transmission of arboviruses: reexamination of and new insights into components, mechanisms, and unique traits as well as their evolutionary trends. Clin Microbiol Rev. 2005;18(4):608–37. Epub 2005/10/15. doi: 10.1128/cmr.18.4.608-637.2005. PubMed PMID: 16223950; PubMed Central PMCID: PMCPMC1265912.

## References

1. Eastwood G, Sang RC, Guerbois M, Taracha ELN, Weaver SC. Enzootic Circulation of Chikungunya Virus in East Africa: Serological Evidence in Non-human Kenyan Primates. Am J Trop Med Hyg. 2017;97(5):1399–404. Epub 2017/10/11. doi: 10.4269/ajtmh.17-0126. PubMed PMID: 29016323; PubMed Central PMCID: PMCPMC5817753.

2. Raulino R, Thaurignac G, Butel C, Villabona-Arenas CJ, Foe T, Loul S, et al. Multiplex detection of antibodies to Chikungunya, O’nyong-nyong, Zika, Dengue, West Nile and Usutu viruses in diverse non-human primate species from Cameroon and the Democratic Republic of Congo. PLoS Negl Trop Dis. 2021;15(1):e0009028. Epub 2021/01/22. doi: 10.1371/journal.pntd.0009028. PubMed PMID: 33476338; PubMed Central PMCID: PMCPMC7853492.

3. Haddow AJ. Field and laboratory studies on an African monkey, Cercopithecus ascanius schmidti Matschie. Proc Zoo Soc Lon. 1952;122(2):297–394. doi: 10.1111/j.1096-3642.1952.tb00316.x.

4. McCrae AW, Henderson BE, Kirya BG, Sempala SD. Chikungunya virus in the Entebbe area of Uganda: isolations and epidemiology. Trans R Soc Trop Med Hyg. 1971;65(2):152–68. Epub 1971/01/01. doi: 10.1016/0035-9203(71)90212-4. PubMed PMID: 4997499.

5. Diallo M, Thonnon J, Traore-Lamizana M, Fontenille D. Vectors of Chikungunya virus in Senegal: current data and transmission cycles. Am J Trop Med Hyg. 1999;60(2):281–6. Epub 1999/03/11. doi: 10.4269/ajtmh.1999.60.281. PubMed PMID: 10072152.

6. McIntosh BM, Paterson HE, McGillivray G, Desousa J. Further Studies on the Chikungunya Outbreak in Southern Rhodesia in 1962. Mosquitoes, Wild Primates and Birds in Relation to the Epidemic. Ann Trop Med Parasitol. 1964;58:45–51. Epub 1964/03/01. doi: 10.1080/00034983.1964.11686213. PubMed PMID: 14147664.

7. Kaschula VR, Van Dellen AF, de Vos V. Some infectious diseases of wild vervet monkeys (Cercopithecus aethiops pygerythrus) in South Africa. J S Afr Vet Assoc. 1978;49(3):223–7. Epub 1978/09/01. PubMed PMID: 218005.

8. McIntosh BM. Antibody against Chikungunya virus in wild primates in Southern Africa. S Afr J Med Sci. 1970;35(3):65–74. Epub 1970/12/01. PubMed PMID: 4398581.

9. Sow A, Faye O, Diallo M, Diallo D, Chen R, Faye O, et al. Chikungunya Outbreak in Kedougou, Southeastern Senegal in 2009-2010. Open Forum Infect Dis. 2018;5(1):ofx259. Epub 2018/01/23. doi: 10.1093/ofid/ofx259. PubMed PMID: 29354659; PubMed Central PMCID: PMCPMC5767945.

10. Inoue S, Morita K, Matias RR, Tuplano JV, Resuello RR, Candelario JR, et al. Distribution of three arbovirus antibodies among monkeys (Macaca fascicularis) in the Philippines. J Med Primatol. 2003;32(2):89–94. Epub 2003/06/26. doi: 10.1034/j.1600-0684.2003.00015.x. PubMed PMID: 12823631.

11. Vourc’h G, Halos L, Desvars A, Boué F, Pascal M, Lecollinet S, et al. Chikungunya antibodies detected in non-human primates and rats in three Indian Ocean islands after the 2006 ChikV outbreak. Vet Res. 2014;45(1):52. Epub 2014/06/03. doi: 10.1186/1297-9716-45-52. PubMed PMID: 24885529; PubMed Central PMCID: PMCPMC4018978.

12. Suhana O, Nazni WA, Apandi Y, Farah H, Lee HL, Sofian-Azirun M. Insight into the origin of chikungunya virus in Malaysian non-human primates via sequence analysis. Heliyon. 2019;5(12):e02682. Epub 2019/12/24. doi: 10.1016/j.heliyon.2019.e02682. PubMed PMID: 31867449; PubMed Central PMCID: PMCPMC6906679.

13. Sam IC, Chua CL, Rovie-Ryan JJ, Fu JY, Tong C, Sitam FT, et al. Chikungunya Virus in Macaques, Malaysia. Emerg Infect Dis. 2015;21(9):1683–5. Epub 2015/08/21. doi: 10.3201/eid2109.150439. PubMed PMID: 26291585; PubMed Central PMCID: PMCPMC4550141.

14. Tongthainan D, Mongkol N, Jiamsomboon K, Suthisawat S, Sanyathitiseree P, Sukmak M, et al. Seroprevalence of Dengue, Zika, and Chikungunya Viruses in Wild Monkeys in Thailand. Am J Trop Med Hyg. 2020;103(3):1228–33. Epub 2020/06/27. doi: 10.4269/ajtmh.20-0057. PubMed PMID: 32588813; PubMed Central PMCID: PMCPMC7470562.

15. Marchette NJ, Rudnick A, Garcia R, MacVean DW. Alphaviruses in Peninusular Malaysia: I. Virus isolations and animal serology. Southeast Asian J Trop Med Public Health. 1978;9(3):317–29. Epub 1978/09/01. PubMed PMID: 34888.

16. Ha DQ, Calisher CH, Tien PH, Karabatsos N, Gubler DJ. Isolation of a newly recognized alphavirus from mosquitoes in Vietnam and evidence for human infection and disease. Am J Trop Med Hyg. 1995;53(1):100–4. Epub 1995/07/01. PubMed PMID: 7625527.

17. Harrison VR, Marshall JD, Guilloud NB. The presence of antibody to Chikungunya and other serologically related viruses in the sera of sub-human primate imports to the United States. J Immunol. 1967;98(5):979–81. Epub 1967/05/01. PubMed PMID: 4960851.

18. Evans TS, Aung O, Cords O, Coffey LL, Wong T, Weiss CM, et al. Sylvatic Transmission of Chikungunya Virus among Nonhuman Primates in Myanmar. Emerg Infect Dis. 2022;28(12):2548–51. Epub 2022/11/24. doi: 10.3201/eid2812.220893. PubMed PMID: 36417997; PubMed Central PMCID: PMCPMC9707571.

19. Nakgoi K, Nitatpattana N, Wajjwalku W, Pongsopawijit P, Kaewchot S, Yoksan S, et al. Dengue, Japanese encephalitis and Chikungunya virus antibody prevalence among captive monkey (Macaca nemestrina) colonies of Northern Thailand. Am J Primatol. 2014;76(1):97–102. Epub 2013/10/10. doi: 10.1002/ajp.22213. PubMed PMID: 24105916.

20. Kading RC, Borland EM, Cranfield M, Powers AM. Prevalence of antibodies to alphaviruses and flaviviruses in free-ranging game animals and nonhuman primates in the greater Congo basin. J Wildl Dis. 2013;49(3):587–99. Epub 2013/06/20. doi: 10.7589/2012-08-212. PubMed PMID: 23778608.

21. Moreira-Soto A, Carneiro IO, Fischer C, Feldmann M, Kummerer BM, Silva NS, et al. Limited Evidence for Infection of Urban and Peri-urban Nonhuman Primates with Zika and Chikungunya Viruses in Brazil. mSphere. 2018;3(1). Epub 2018/02/07. doi: 10.1128/mSphere.00523-17. PubMed PMID: 29404420; PubMed Central PMCID: PMCPMC5793042.

22. Laroque PO, Valença-Montenegro MM, Ferreira DRA, Chiang JO, Cordeiro MT, Vasconcelos PFC, et al. Levantamento soroepidemiológico para arbovírus em macaco-prego-galego (Cebus flavius) de vida livre no estado da Paraíba e em macaco-prego (Cebus libidinosus) de cativeiro do nordeste do Brasil. Pesq Vet Bras. 2014;34:462–8.

23. Batista PM, Andreotti R, Chiang JO, Ferreira MS, Vasconcelos PF. Seroepidemiological monitoring in sentinel animals and vectors as part of arbovirus surveillance in the state of Mato Grosso do Sul, Brazil. Rev Soc Bras Med Trop. 2012;45(2):168–73. Epub 2012/04/27. doi: 10.1590/s0037-86822012000200006. PubMed PMID: 22534986.

24. Perez JG, Carrera JP, Serrano E, Pitti Y, Maguina JL, Mentaberre G, et al. Serologic evidence of zoonotic alphaviruses in humans from an indigenous community in the Peruvian Amazon. Am J Trop Med Hyg. 2019;101(6):1212–18. Epub 2019/10/02. doi: 10.4269/ajtmh.18-0850. PubMed PMID: 31571566.

25. de Thoisy B, Gardon J, Salas RA, Morvan J, Kazanji M. Mayaro virus in wild mammals, French Guiana. Emerg Infect Dis. 2003;9(10):1326–9. Epub 2003/11/12. doi: 10.3201/eid0910.030161. PubMed PMID: 14609474; PubMed Central PMCID: PMCPMC3033094.

26. Hoch AL, Peterson NE, LeDuc JW, Pinheiro FP. An outbreak of Mayaro virus disease in Belterra, Brazil. III. Entomological and ecological studies. Am J Trop Med Hyg. 1981;30(3):689–98. Epub 1981/05/01. doi: 10.4269/ajtmh.1981.30.689. PubMed PMID: 6266265.

27. Díaz LA, del Pilar Díaz M, Almirón WR, Contigiani MS. Infection by UNA virus (Alphavirus; Togaviridae) and risk factor analysis in black howler monkeys (Alouatta caraya) from Paraguay and Argentina. Transactions of the Royal Society of Tropical Medicine and Hygiene. 2007;101(10):1039–41. doi: 10.1016/j.trstmh.2007.04.009.

28. Seymour C, Peralta PH, Montgomery GG. Serologic evidence of natural togavirus infections in Panamanian sloths and other vertebrates. Am J Trop Med Hyg. 1983;32(4):854–61. Epub 1983/07/01. doi: 10.4269/ajtmh.1983.32.854. PubMed PMID: 6309027.

29. Degallier N, Travassos da Rosa AP, Vasconcelos PFC, Hervé JP, Sa Filho GC, Travassos da Rosa JFS, et al. Modifications of arbovirus transmission in relation to construction of dams in Brazilian Amazonia. Ciência e Cultura. 1992;44:124–35.

